# Cryptic transmission of SARS-CoV-2 and the first COVID-19 wave in Europe and the United States

**DOI:** 10.1101/2021.03.24.21254199

**Authors:** Jessica T. Davis, Matteo Chinazzi, Nicola Perra, Kunpeng Mu, Ana Pastore y Piontti, Marco Ajelli, Natalie E. Dean, Corrado Gioannini, Maria Litvinova, Stefano Merler, Luca Rossi, Kaiyuan Sun, Xinyue Xiong, M. Elizabeth Halloran, Ira M. Longini, Cécile Viboud, Alessandro Vespignani

## Abstract

Given the narrowness of the initial testing criteria, the SARS-CoV-2 virus spread through cryptic transmission in January and February, setting the stage for the epidemic wave experienced in March and April, 2020. We use a global metapopulation epidemic model to provide a mechanistic understanding of the global dynamic underlying the establishment of the COVID-19 pandemic in Europe and the United States (US). The model is calibrated on international case introductions at the early stage of the pandemic. We find that widespread community transmission of SARS-CoV-2 was likely in several areas of Europe and the US by January 2020, and estimate that by early March, only 1 − 3 in 100 SARS-CoV-2 infections were detected by surveillance systems. Modeling results indicate international travel as the key driver of the introduction of SARS-CoV-2 with possible importation and transmission events as early as December, 2019. We characterize the resulting heterogeneous spatio-temporal spread of SARS-CoV-2 and the burden of the first COVID-19 wave (February-July 2020). We estimate infection attack rates ranging from 0.78%-15.2% in the US and 0.19%-13.2% in Europe. The spatial modeling of SARS-CoV-2 introductions and spreading provides insights into the design of innovative, model-driven surveillance systems and preparedness plans that have a broader initial capacity and indication for testing.

## Introduction

The first confirmed case of COVID-19 in the United States (US) was diagnosed in Washington state on January 21, 2020 (1). In Europe, the first three COVID-19 cases were reported in France on January 24 and had an onset of symptoms on January 17, 19 and 23, respectively (2; 3). In quick succession other cases were confirmed in the US (4; 5; 6) and in several European countries such as Germany (January 27), Italy (January 30), Spain, and the United Kingdom (January 31). In Fig. 1A we include a timeline of initial confirmed cases and early containment and mitigation initiatives in the US and Europe. To study the cryptic spreading phase and the ensuing first wave of the COVID-19 pandemic, we use a data-driven, stochastic, spatial, and age-structured global epidemic model. We develop a mechanistic understanding of the epidemic evolution and estimate the time frame for the establishment of local transmission in different states and countries. The model provides a statistical picture of SARS-CoV-2 introductions across US states and European countries. We quantify the association between the amount of international/domestic air travel and the model estimates of the arrival times of the virus in the US and Europe, showing that international and domestic travel patterns were a key driver in the establishment of SARS-CoV-2 local transmission. Furthermore, we use the model to estimate the COVID-19 disease burden across the two regions. We provide model estimates for the infection fatality ratios and the infection attack rates as of July 4, 2020. We find that our model-estimated infection attack rates are in good agreement with the results from serological studies of SARS-CoV-2 antibody prevalence conducted at different spatial resolutions (i.e., city, state, country). Additionally, the model highlights a strong statistical association between the number of cases reported at the time of issuing major mitigation policies in each country/state and the estimated number of infections at the end of the first wave. This is in agreement with statistical analysis showing that the effectiveness of mitigation policies is associated with the timing of their adoption (7; 8; 9; 10; 11; 12; 13; 14; 15). The unique, mechanistic understanding of the way in which the COVID-19 pandemic unfolded highlights that, in countries with confirmed local transmission, policies such as testing based on travel history and international travel restrictions are highly inefficient in preventing the development of local outbreaks. Wide spread testing would have detected transmission earlier and allowed for earlier implementation of interventions. Future preparedness plans must have broader, initial capacity and indication for testing to mitigate wide spread cryptic transmission. These findings are of particular relevance given that contrasting the spread of SARS-CoV-2 variants of concern presents similar dynamics and problems.

**Figure 1:**
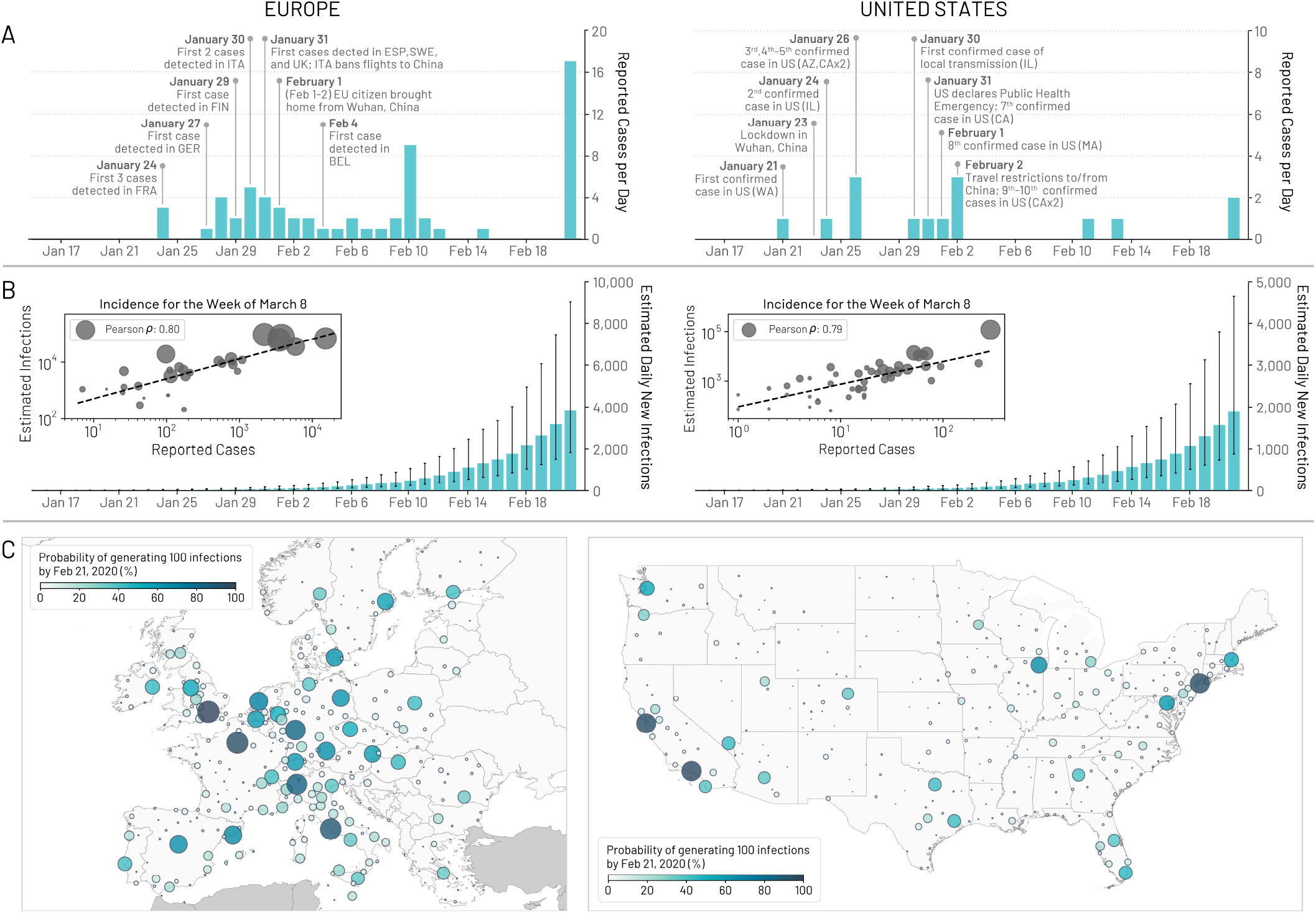
Early picture of the COVID-19 outbreak in Europe and the United States. (A) Timelines of the daily reported and confirmed cases of COVID-19 in Europe and US including information on initial reported cases and other significant events related to the outbreak. (B) Model-based estimates for the daily number of new infections in Europe and US. The inset plot compares the weekly incidence of reported cases with the weekly incidence of infections estimated by the model for the week of March 8 − 14, 2020 for the continental US-states and European countries that reported at least 1 case. Circle size corresponds to the population size of each state and country. (C) The probability that a city in Europe and the US had generated at least 100 cumulative infections by February 21, 2020. Color and circle size are proportional to the probability.

## Results

We consider data concerning the continental US and the following list of 30 countries we will informally refer to as “Europe”: Austria, Belgium, Bulgaria, Croatia, Czech Republic, Denmark, Estonia, Finland, France, Germany, Greece, Hungary, Iceland, Ireland, Italy, Latvia, Lithuania, Luxembourg, Malta, Netherlands, Norway, Poland, Portugal, Romania, Slovak Republic, Slovenia, Spain, Sweden, Switzerland, and the UK. To study the spatial and temporal spread of SARS-CoV-2, we use the Global Epidemic and Mobility Model (GLEAM), a stochastic, spatial, and age-structured metapopulation epidemic model (16; 17; 18). The model was previously used to characterize the early stage of the COVID-19 epidemic in mainland China and the effect of travel restrictions on infections exported to other regions (19). The model divides the global population into more than 3, 200 subpopulations in roughly 200 different countries and territories. A subpopulation is defined as the catchment area around major transportation hubs. The airline transportation data encompass daily origin-destination traffic flows from the Official Aviation Guide (OAG) database (20) reflecting actual traffic changes that occurred during the pandemic. Ground mobility and commuting flows are derived from the analysis and modeling of data collected from the statistics offices of 30 countries on five continents (17; 16). We set, as initial conditions, an epidemic starting date in Wuhan, China between November 15, 2019 and December 1, 2019, with 20 initial infections (21; 22; 23; 24; 19; 25). This considers that the virus could have emerged as early as mid October, 2019. The international travel data account for travel restrictions and government issued policies. Furthermore, the model accounts for the reduction of internal, country-wide mobility and changes in contact patterns in each country and state in 2020. To initially calibrate the global model, we use an Approximate Bayesian computation (ABC) method (26) that considers reports of international travelers carrying SARS-CoV-2 returning from China up to January 21, 2020, and accounts for different case detection capabilities (27; 19) among countries. The model details are reported in the Materials and Methods section and the Supplementary Information (SI).

Stochastic simulations of the global epidemic spread yield international and domestic infection importations, incidence of infections, and deaths per subpopulation at a daily resolution. In Fig. 1B we show the model estimates for the median daily incidence of new infections up to February 21, 2020, for both the US and Europe. These values are much larger than the number of officially reported cases (see Fig.1A), highlighting the significant number of potential transmission events that may have already occurred before many states and countries had implemented testing strategies independent of travel history.

As validation we compare our model projections of the number of infections during the week of March 8, 2020 to the number of cases reported during that week within each US state and European country that had at least 1 reported case (shown in Fig.1B inset). While we see a strong correlation between the reported cases and our model’s projected number of infections (Pearson’s correlation coefficient on log-values, US: 0.79, *p <* 0.001; Europe: 0.80, *p <* 0.001), many fewer cases had actually been reported by that time. If we assume that the number of reported cases and simulated infections are related through a simple binomial sampling process, we find that on average 9 in 1, 000 infections (90%CI [1 – 35 per 1, 000]) and 35 in 1, 000 infections (90%CI [4 – 90 per 1, 000]) were detected by March 8, 2020 in the US and Europe respectively. As testing capacity increased that week, the ascertainment rate grows and our estimates increase to detecting 17 in 1, 000 infections (90%CI [2 – 55 per 1, 000]) by March 14, 2020 in the US and 77 in 1, 000 infections (90%CI [5 – 166 per 1, 000]) in Europe. The estimated ascertainment rates are in agreement with independent results based on different statistical methodologies (28; 29; 30). In mid-February, other than travel-related restrictions, there were very few mitigation policies implemented for the purpose of reducing community transmission (i.e., social distancing guidelines, school closures, stay at home orders, etc.) in the US and Europe. Combined with the lack of testing capabilities, the virus was able to spread undetected and unhindered. In Fig.1C we show the probability that a city in the US or Europe had generated at least 100 infections by February 21, 2020. We see that the progression of the virus through the US and Europe is both temporally and spatially heterogeneous. While many cities had not yet experienced much community transmission by late February, a few areas such as New York City or London likely already had local virus spreading. As discussed in more detail in the following sections, the position of the cities within the global mobility network plays a critical role in the timing of the virus’ introduction to the population and onset of local transmission.

### Onset of local transmission

The model allows us to study the unfolding patterns compatible with the global importation of cases from China leading to the initial local outbreaks. It is important to stress that the model’s realizations explore all possible paths of the epidemic. Thus rather than describing a specific, single causal chain of events, the results provide a statistical description of all the potential pandemic histories compatible with the initial evolution of the pandemic in China. For instance, some initial clusters, such as in Germany, have been effectively contained, possibly delaying the start of wide spread transmission (31). While the inclusion of these additional events in selecting epidemic paths would be computationally unfeasible, it is possible to assume, considering also recent evidence from models of genomic epidemiology (32; 31; 33), that Italy has been the first among European countries to experience substantial widespread transmission. Due to limited testing capacity during the early phases of the pandemic, confirmed SARS-CoV-2 deaths might be a better proxy for the relative start of the local outbreak in each country rather than reported cases. As of March 9, 2020, Italy reported 463 cumulative deaths, Spain 35, USA 26, France 25, Germany 2, the UK 7, and Italy was the first in the region to impose a national lockdown. Therefore, throughout the paper, we constrain the ensemble of simulations focusing only on stochastic realizations where Italy is the first country, in the group under examination, to experience sustained local transmission. In the SI we report the analysis of the full unconstrained set of simulations.

Here, we define the onset of local transmission for a country or state as the earliest date when at least 10 new infections are generated per day. This number is chosen because at this threshold the likelihood of stochastic extinction is extremely small (34; 35). In fact, the probability of disease extinction in a fully susceptible and well-mixed population exposed to *n* infected individuals can be approximated as 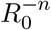, which is approximately 10^*−*5^ when *R*_0_ = 2.7. As detailed in the SI, further calibration on the US states and European countries suggests values of *R*_0_ ranging from 2.4 − 2.8. These values are consistent with many other (country dependent) estimates (36; 37; 38; 39; 40; 8). At the same time, given the doubling time of COVID-19 before the implementation of public health measures, any variation of a factor 2 around the 10 infections/day threshold corresponds to a small adjustment of 3 − 5 days to the presented timelines.

In Fig. 2, we show the posterior distribution of onset of local transmission for different US states (A) and European countries (B). Among the US states, California and New York state are the earliest, with over a 50% estimated probability of local transmission by the end of January (California) or beginning of February (New York), 2020. In Europe, Italy, UK, Germany, and France are the first countries with a probability larger than 50% to have experienced local transmission by the end of January 2020. A majority of the states and countries analyzed have a median date of onset of local transmission by early March, with the large majority of them in February, 2020, a critical month for the cryptic spread of SARS-CoV-2 in the continental US and Europe. Remarkably, the plots for both Europe and the US indicate that while surveillance and testing in February and early March were focused on travel history, several European countries and US states were likely already experiencing local community transmission. This finding confirms that from late January to early March SARS-CoV-2 had been spreading across the US and Europe mostly undetected. However, the wide distribution of dates suggest that SARS-CoV-2 cryptic transmission may have begun as early as December, 2019. The model also allows us to estimate possible COVID-19 related deaths during the undetected spreading phase. For instance by March 1, 2020, the model suggests that the median cumulative number of deaths was 56 [90% CI 9 − 544] in the US and 120 [90% CI 21 − 1, 177] in Europe. Although some US states and European countries launched investigations in search of evidence that COVID-19 was the cause of death as far back as December 2019, it is likely that most early deaths were not recorded (41).

**Figure 2:**
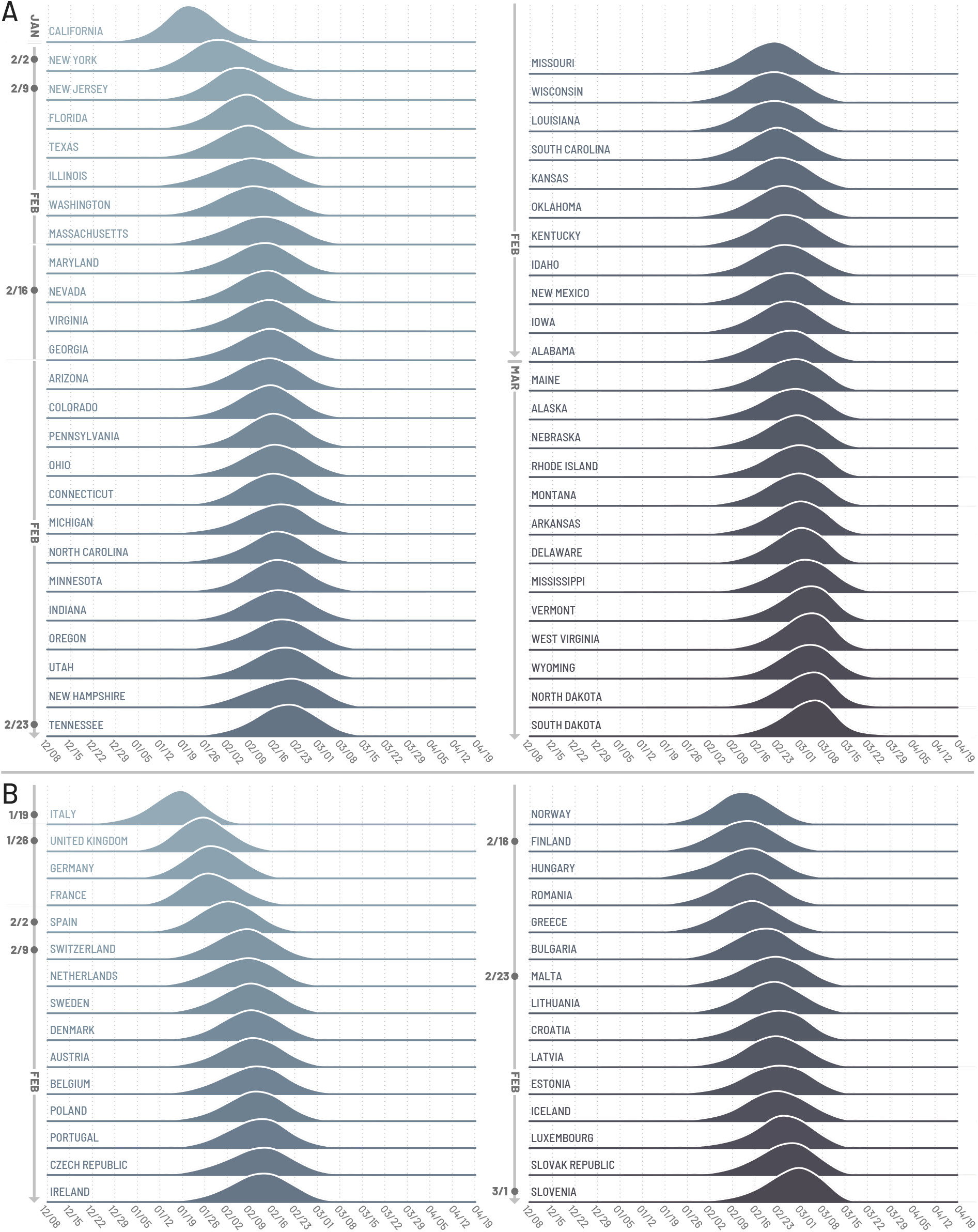
Timing of the onset of local transmission. We plot the posterior distributions of the week when each US state (A) or European country (B) first reached 10 locally generated SARS-CoV-2 transmission events per day. Countries/states are ordered by the median date of their posterior distribution. The week of this date corresponds to the dates reported on the the vertical axis.

### SARS-CoV-2 introductions

As the model allows the recording of the origin and destination of SARS-CoV-2 carriers at the global scale, we can study the possible sources of infection importation for each US state and European country. More specifically, we record the number of introductions in each stochastic realization of the model. In Fig. 3 we visualize the origin of the introductions considering some key geographical regions (e.g., Europe and Asia) while keeping the US and China separate and aggregating all the other countries (i.e., Others). We show the directed importation flows from the aggregated source regions to the US states (A) and European countries (B). States and countries are ordered according to the date of the estimated establishment of local transmission. In both cases, the contribution from mainland China is barely visible and the local share (i.e., sources within Europe and US) becomes significantly higher across the board. Hence, while importation events in the early phases of the outbreak were key to start the local spreading (see details in the SI), the cryptic transmission phase has been sustained largely by internal flows. Domestic SARS-CoV-2 introductions through April 30, 2020, account for 71% [IQR 61% − 82%] of the introductions in California, 79% [IQR 73% − 88%] in Texas, and 71% [IQR 61% − 82%] in Massachusetts. European origins account for 69% [IQR 60% − 80%], 84% [IQR 79% − 91%], and 58% [IQR 48% − 68%] of the introductions in Italy, Spain, and the UK, respectively. In the SI we report the full breakdown for all states and countries.

**Figure 3:**
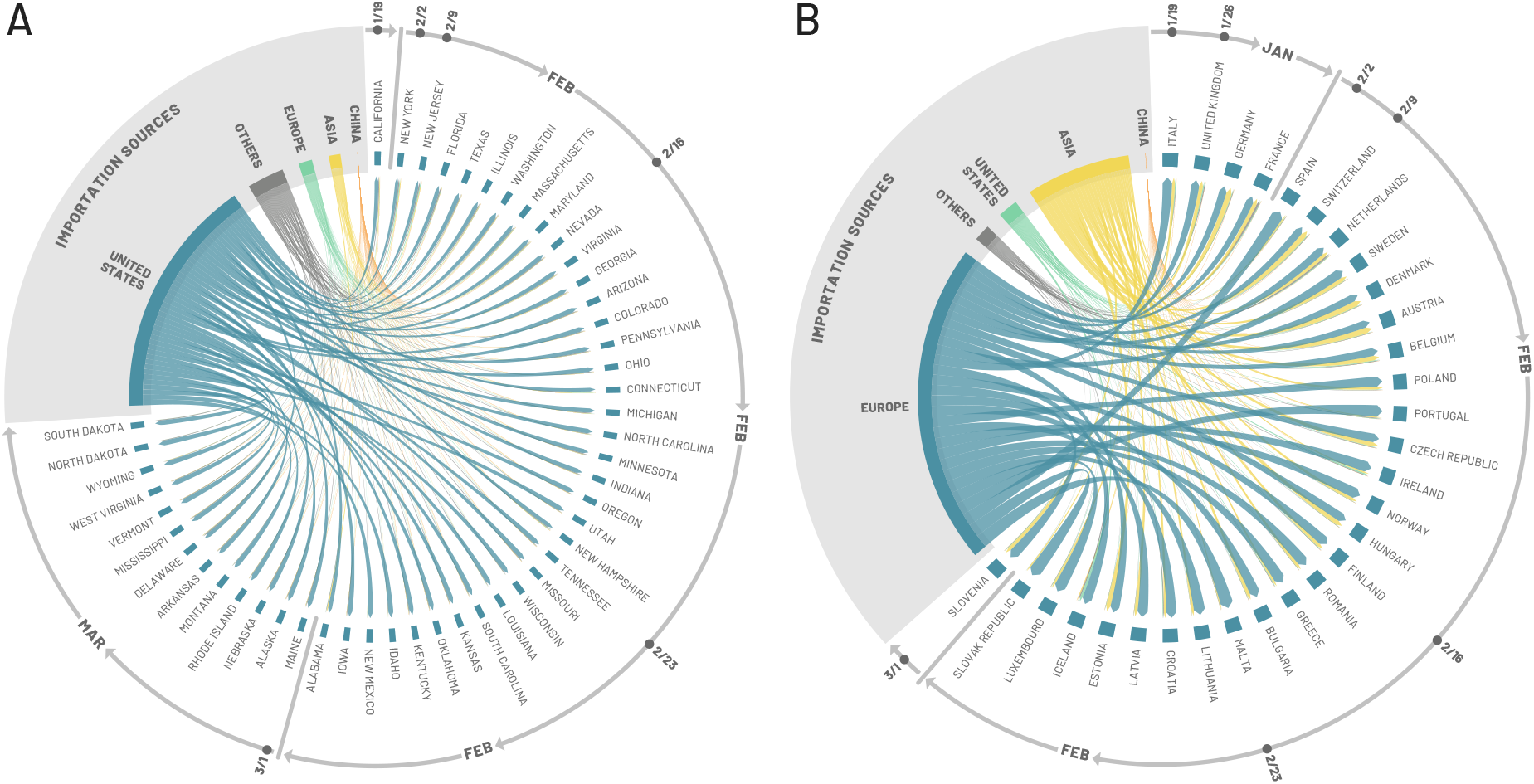
Importation sources. Each US state (A) and European country (B) is displayed in a clockwise order with respect to the start of the local outbreak (as seen in Fig. 2). Importation flows are directed and weighted. We normalize links considering the total in-flow for each state so that the sum of importations flows, for each state, is one. In the SM we report the complete list of countries contributing, as importation sources, in each group (i.e., geographical region).

It is important to distinguish between the full volume of SARS-CoV-2 introductions and the introduction events that could be relevant to the early onset of local transmission in each stochastic realization of the model. In the model we can investigate these specific events by recording introduction events before the local transmission chains were established (defined as the median dates of Fig. 2). We report the results of this analysis in the SI, showing that importations from mainland China may be relevant in seeding the epidemic in January, but then play a small role in the COVID-19 expansion in the US and Europe due to the travel restrictions imposed to/from mainland China after January 23, 2020. In fact, about 74% [IQR 60% − 100%] and 45% [IQR 15% − 71%] of the virus introduction before the onset of local transmission in California and New York states, respectively, were from mainland China. The equivalent share of importations from China in Italy and the UK were 72% [IQR 50% − 100%] and 52% [IQR 31% − 71%].

Our results concerning SARS-CoV-2 introductions in different countries/states can be compared to analysis based on gene sequencing and travel volumes, showing a good degree of agreement with the temporal and geographical distribution of SARS-CoV-2 importations. For example, Ref. (42) estimates that the majority of importation events associated with onward transmissions in the UK, through April 2020, came from Europe. Similar to our findings, the contributions from China are quantified below 1% and limited to the very early phase. Furthermore, the seeding events from the US are estimated to be ∼ 3% which is in agreement with our estimate of 8% [IQR 3% − 9%]. However, their results point to a larger share from Europe (∼ 90%) compared to ours (58% [IQR 48% − 68%]). Conversely, we estimate a larger contribution from Asia (27% [IQR 19% − 35%]). The discrepancies might be due to biases in genomic sampling (43) and/or the fact that we sample all possible epidemic paths statistically possible rather than the single, observed occurrence. To this point, it is worth stressing that seeding importations are different from the actual number of times the virus has been introduced to each location with subsequent onward transmission. Even after local transmission has started, future importation events may give rise to additional onward transmission forming independently introduced transmission lineages of the virus(42). Ref. (44) confirms the key role of national importations in the US. In fact, by analyzing both genomic and travel data (national and international) it estimates that the outbreak in Connecticut was largely driven by domestic importations. Our results indicate an internal share of importations for that state of 64% [IQR 54% − 76%]. Furthermore, Ref. (33) confirms the key role of importations from China to Italy at the beginning of the pandemic. As shown in the SI and mentioned above, our results suggest that 72% of the early introductions before the start of the local outbreak in Italy are linked to China.

### COVID-19 burden

The model allows us to estimate the disease burden in the US and Europe once COVID-19 has established local transmission. Starting in March 2020, the COVID-19 epidemic trajectory in each country and state is driven by the establishment and timing of non-pharmaceutical interventions (NPIs) as well as by the epidemiological relevant features (i.e., population size and density, age-structure etc.) which are spatially heterogeneous (45; 46; 47; 7). The model accounts for these features as detailed in the SI. To calibrate the model results for each individual US state and European country, we use the weekly number of new deaths reported from March 22, 2020 to June 27, 2020. Furthermore, we consider a uniform prior for the average infection fatality ratio (IFR) in the range from 0.4% to 2% that is age stratified proportional to the IFR values reported in Ref. (48). We also consider a uniform prior for reporting delays between the date of death and reporting ranging from 2 to 22 days in both Europe and the US (49). We provide the details of our calibration for each geographical unit (i.e., US state or European country) in the SI.

In Fig. 4(A-D, F-I), we report the projected results of the weekly deaths of the first wave for selected states and countries. Additional model results for all investigated regions including a sensitivity analysis of different calibration methods can be found in the SI. We find a strong correlation between the weekly model-estimated deaths and the reported values with a Pearson correlation coefficient of 0.99 (*p <* 0.001) for both Europe and the US (see Fig. S5). As the data suggest, many European countries and US states saw peaks in April and May with various decreasing trajectories that are dependent on the mitigation strategies in place. Additionally, we report the estimated cumulative infection attack rates and IFRs as of July 4, 2020, in European countries experiencing more than 100 total deaths and the top 20 states ranked by infection attack rate in the US. The median infection attack rates vary from 0.19% − 13.2% in Europe and 0.78% − 15.2% in the US.

**Figure 4:**
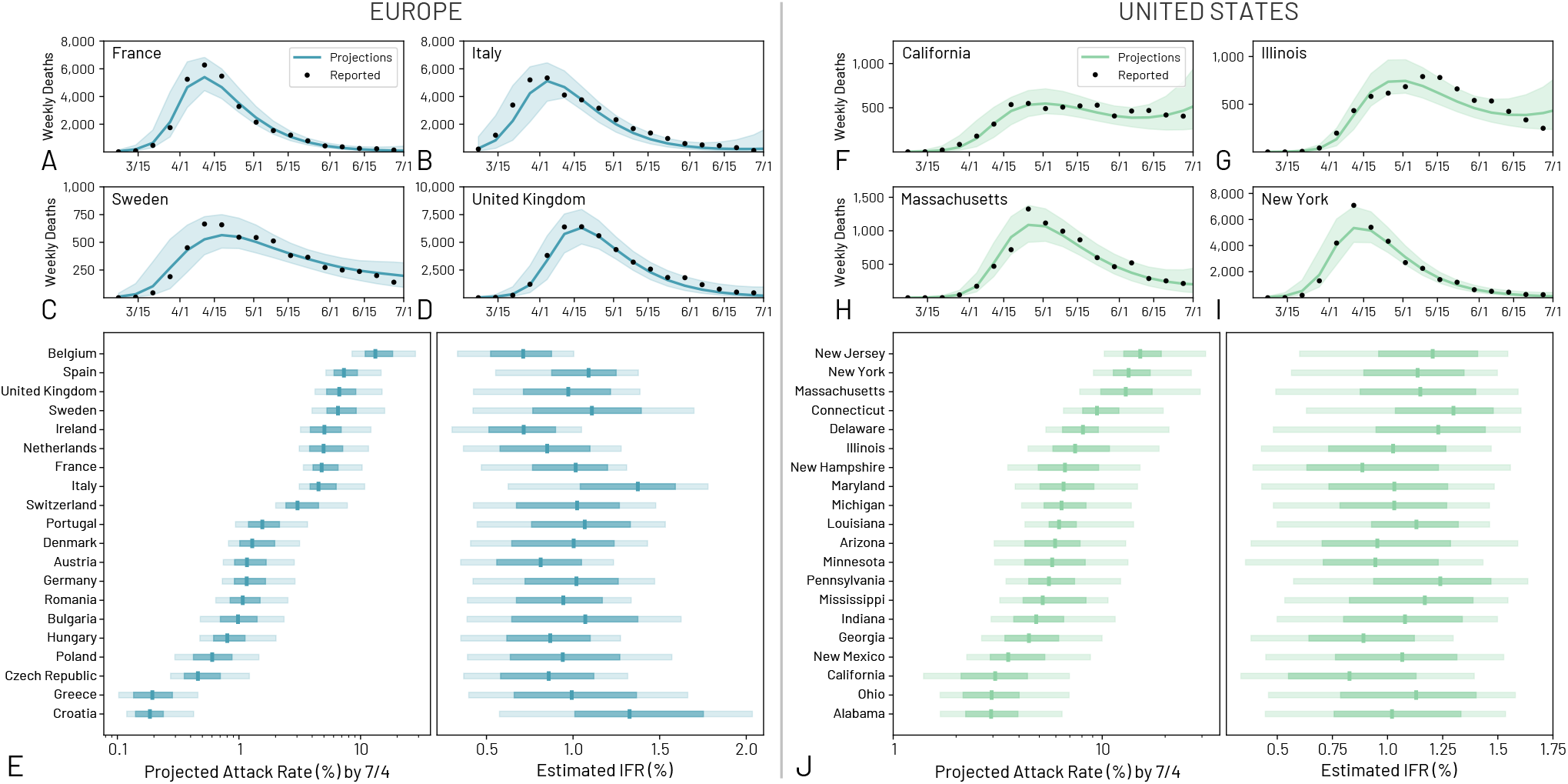
The burden of the first wave in Europe and the US. (A-D) Model projection results of the weekly deaths for selected countries in Europe. (E) Estimated infection attack rates and infection fatality rates by July 4, 2020 for European countries where there were at least 100 reported deaths. (F-I) Model projection results of the weekly deaths for selected states in the US. (J) Estimated infection attack rates and infection fatality rates by July 4, 2020 for 20 US states.

Within Europe, Belgium has the highest estimated infection attack rate of 13.2% (90% CI [8.5% − 28.3%]) by July 4, 2020, in agreement with the results detailed in Ref. (28). Furthermore, by that time Belgium reported the highest COVID-19 mortality rate out of the European countries investigated with 8.5 deaths per 10, 000 individuals. Italy is estimated to have the highest median IFR of 1.4% (90% CI [0.6% − 1.8%]), which aligns with other ranges reported in the literature (50; 51). The US states with the highest infection attack rates are located within the Northeast and experienced a significant first wave during March-April 2020. New York and New Jersey are the top two states with infection attack rates of 13.4% (90% CI [9.1% − 26.7%]) and 15.2% (90% CI [10.2% − 31.3%]) respectively. These numbers are aligned with estimates from New York City reported in Ref. (52). However, unlike many European countries, some states did not experience a significant initial wave until late summer 2020, after the time window considered here. The IFRs estimated for the US states range from 0.8% to 1.3%. In the SI we report summary tables with estimated IFRs, infection attack rates, as well as the reproductive number in the absence of mitigation measures for all calibrated US states and European countries.

### The drivers and impact of the cryptic transmission phase

In the early stages of a pandemic, surveillance data are known to be unreliable due to under-detection. For each state in the US and each country in Europe we compared the order in which they surpassed 100 cumulative infections in the model and in total cases in the surveillance data (gathered from the John Hopkins University Coronavirus Resource Center (53)). In Fig. 5A we plot the ordering for states and compute the Kendall rank correlation coefficient *τ* (see SI for details). The correlation is positive (*τ*_*EU*_ = 0.71, *p <* 0.001 and *τ*_*US*_ = 0.68, *p <* 0.001) indicating that, despite the detection and testing issues, the expected patterns of epidemic diffusion are largely described by the model in both regions. The model, however, suggests that one major driver of this early diffusion pattern is air travel. We compare the ordering of states and countries according to their air travel volume to their epidemic order as previously defined (Fig. 5A). We consider both national and international traffic, and find a positive correlation (*τ*_*EU*_ = 0.66 with *p <* 0.001 and *τ*_*US*_ = 0.66 with *p <* 0.001) between the epidemic ordering derived from surveillance data and air traffic, suggesting the passenger volume of both international and national traffic are key factors driving the early spreading of the outbreak across countries. Similar observations have been reported in China, where the initial spreading of the virus outside Hubei was strongly correlated with the traffic to/from the province (54). Population size is also correlated with both the traveling flows (*τ*_*EU*_ = 0.59, *p <* 0.001 and *τ*_*US*_ = 0.7, *p <* 0.001) and the epidemic order of each state (*τ*_*EU*_ = 0.46, *p <* 0.001 and *τ*_*US*_ = 0.68, *p <* 0.001) as discussed in the SI. In our model, it is not possible to exclude increased contacts in highly populated places before social distancing interventions and disentangle this effect from increased seeding due to the correlation between travel volume and population size.

**Figure 5:**
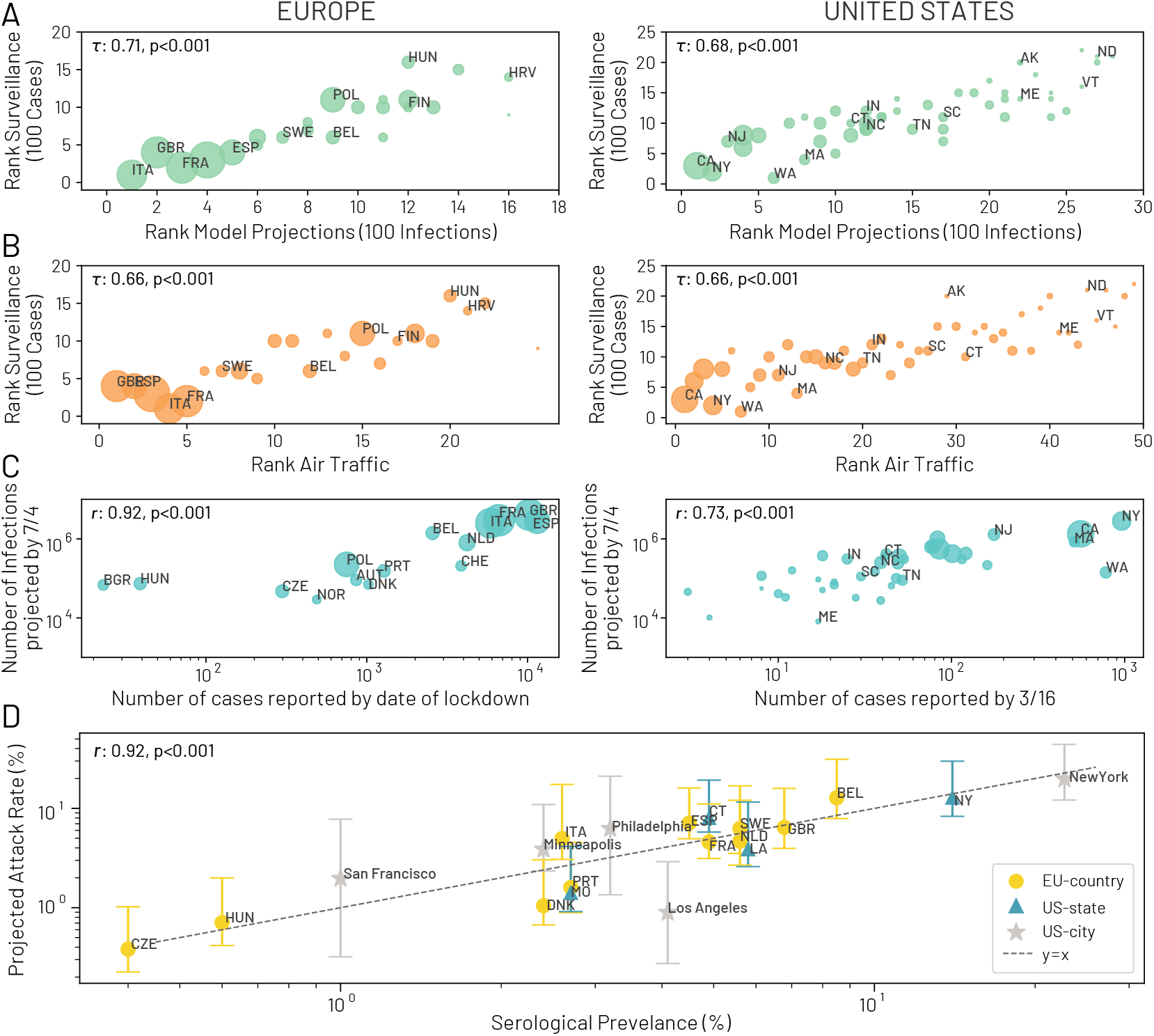
Correlation Analysis for European countries and US states. (A) The correlation between the ordering of each country/state to reach 100 infections in the model projections and to reach 100 reported cases in the surveillance data. Correlation is computed considering the Kendall rank correlation coefficient, *τ*. (B) The correlation between the ordering of each country/state considering the time needed to reach 100 reported cases in the surveillance data and the ranking of the combined international and domestic air traffic. (C) Left: the correlation between the number of cases reported by the date of lockdown for selected European countries (from Table 4 in (55)) and the projected total number of infections by July 4, 2020. Right: the correlation between the number of cases reported by March 16, 2020 for each US state and the projected total infections by July 4, 2020. (D) The correlation between the model-projected infection attack rate and the serological prevalence collected from studies. Data points refer to different dates and locations (table with values and dates reported in the SI). The correlations are calculated using the Pearson correlation coefficient *r* in (C-D).

As more cases were detected in countries and states, travel restrictions were implemented to/from high risk regions. Consequently, COVID-19 deaths started to rise along with hospitalizations and many governments began to issue social distancing guidelines, school closures, and community / country-wide lock downs. All NPIs were aimed at mitigating the spread of COVID-19 (8). In Fig. 5C (left) we report the correlation between the cumulative infections projected by the model on July 4, 2020, and the number of cases reported by the date of lockdown (reported data from Ref. (55)). Similarly, in the US (right: Fig. 5C) we show the correlation between the cumulative infection projections on July 4, 2020, and the number of cases reported by March 16, 2020, the date the “15 days to slow the spread” guidelines were released (56). At this point in time, many people were aware of the virus and altering their behavior (such as working from home or social distancing) (57; 58). In both cases we find a strong correlation (Pearson correlation coefficient, *r* = 0.92, *p <* 0.001 and *r* = 0.73, *p <* 0.001 in Europe and US respectively) indicating that the earlier NPIs had been issued with respect to the number of cases confirmed in each specific state or country, the smaller the COVID-19 burden experienced during the first wave. This is in agreement with other analyses showing that the timing of NPIs is crucial in limiting the burden of COVID-19 (7; 8; 9; 10; 11; 12; 13; 14; 15).

By April-May 2020, after a month of *stay-at-home* orders and other NPIs coupled with travel restrictions, many European countries and US states started to observed a decline in SARS-CoV-2 cases, deaths, and hospitalizations. This decline led to the relaxation of many social distancing policies, such as removing *stay-at-home* directives or opening workplaces and restaurants to in-person business. Even though at the end of the first wave the active prevalence of the virus may have been low within these regions, the relative low level of residual immunity left in a population has favored an epidemic resurgence observed both in the US and Europe in October and November 2020. In Fig. 5D we report the correlation between our model-estimated infection attack rates and the prevalence of individuals with the SARS-CoV-2 antibody from 13 serological studies across the US and Europe. We find a remarkably good agreement between model estimates and seroprevalence studies at different point in times. Data from this figure can be found in the Table S7 in the SI.

## Discussion

The model presented here captures the spatial and temporal heterogeneity of the early stage of the pandemic, going beyond the single country-level reconstruction. It provides a mechanistic understanding of the interconnected, underlying dynamics of the pandemic’s evolution. The results of our analysis suggest that the first sustained local transmission chains took place as early as January and by the end of February 2020 the virus was spreading in a majority of European countries and US states. This timeline is shifted several weeks ahead with respect to the detection of cases in surveillance data and is consistent with the fact that, in January and February, no country had the capacity to do mass testing. The results also indicate that the sources of introduction of SARS-CoV-2 infections into Europe and the US changed substantially and rapidly through time. If testing had been more widespread and not restricted to individuals with a travel history from China, there would have been more opportunities for earlier detection and interventions.

The numerical simulations yield posterior distributions characterizing the timing of the onset of local transmission of SARS-CoV-2 across US states and European countries. These results generate a comprehensive picture of the cryptic phase of the pandemic, especially for countries and states where genomic surveillance and testing capacity were not adequate. This is also relevant in the interpretation of many studies that are searching for SARS-CoV-2 traces in existing databases (ex. blood donors, sewage samples etc.).

The model can estimate the impact of the first wave through measuring the country and state level infection attack rates. These projections align with other sources and provide insights into the heterogeneous progression of the pandemic across countries/states due to the timing and magnitude of different public health responses. We find that an early response is important in minimizing the burden of COVID-19 disease on a region. The first European country to implement a *cordon sanitaire* in response to the rise in SARS-CoV-2 related illness was Italy on February 23, 2020, for a few northern cities (59). However, this is nearly one month after the median date of the onset of local transmission estimated by the model (see Fig. 2B). Many other countries followed suit and implemented national lock downs in March 2020 (45; 60), weeks after our model estimated that SARS-CoV-2 was introduced and subsequently spreading.

More generally, our results show that reactive response strategies, such as issuing travel restrictions targeting countries only after local transmission is confirmed, are highly inefficient. These strategies fail to prevent the establishment of local outbreaks because they do not recognize the critical importance of the cryptic phase. In addition, this finding parallels the emerging story of the cryptic spreading of variants of concern, pointing out the need of increased testing capacity not dependent on travel history, contact tracing, and promoting the establishment of gene sequencing surveillance infrastructures.

As with all modeling analyses, results are subject to biases from the limitations and assumptions within the model as well as the data used in its calibration. The model’s parameters, such as generation time, incubation period, and the proportion of asymptomatic infections are chosen according to the current knowledge of SARS-CoV-2. Although the model is robust to variations in these parameters (see the SI for the sensitivity analysis), more information on the key characteristics of the disease would considerably reduce uncertainties. The model calibration does not consider correlations among importations (i.e., family travel) and assumes that travel probabilities are age-specific across all individuals in the catchment area of each transportation hub.

Although the modeling results should be interpreted cautiously in light of the assumptions and limitations inherent to modeling approaches, they are of interest in combination with sequencing data of SARS-CoV-2 genomes to reconstruct in greater detail the early epidemic history of the COVID-19 pandemic. The methods used in this analysis offer a blueprint to identify the most likely early spreading dynamics of emerging variants and they can be used as a real-time risk assessment tool to inform policy makers. Anticipating the locations where the virus is most likely to spread to next could be instrumental in guiding enhanced testing and surveillance activities, and complement phylogeographic inference approaches (33). The estimated SARS-CoV-2 importation patterns and the cryptic transmission phase dynamics are of potential use when planning and developing public health policies in relation to international traveling and they could provide important insights in assessing the potential risk and impact of emerging SARS-CoV-2 variants in regions of the world with limited testing and genomic surveillance resources.

## Methods

### SARS-CoV-2 transmission dynamic

The transmission dynamics take place within each subpopulation and assume a classic SLIR-like compartmentalization scheme for disease progression similar to those used in several large scale models of SARS-CoV-2 transmission (29; 61; 62; 63; 64; 22). Each individual, at any given point in time, is assigned to a compartment corresponding to their particular disease-related state (being, e.g., susceptible, latent, infectious, removed) (19). This state also controls the individual’s ability to travel (details in the SI). Individuals transition between compartments through stochastic chain binomial processes. Susceptible individuals can acquire the virus through contacts with individuals in the infectious category and can subsequently become latent (i.e., infected but not yet able to transmit the infection). The process of infection is modeled using age-stratified contact patterns at the state and country level (65; 66). Latent individuals progress to the infectious stage at a rate inversely proportional to the latent period, and infectious individuals progress to the removed stage at a rate inversely proportional to the infectious period. The sum of the mean latent and infectious periods defines the generation time. Removed individuals are those who can no longer infect others. To estimate the number of deaths, we use as prior the age-stratified infection fatality ratios from Ref (48). The transmission model does not assume heterogeneities due to age differences in susceptibility to the SARS-CoV-2 infection for younger children (1 − 10 years old). This is an intense area of discussion (67; 68; 69).

### Model calibration

We assume a start date of the epidemic in Wuhan, China, that falls between November 15, 2019 and December 1, 2019, with 20 initial infections (21; 22; 23; 24; 19). The model generates an ensemble of possible epidemic realizations and is initially calibrated using Approximate Bayesian computation (ABC) rejection approach (26) based on the observed international importations from mainland China through January 21, 2020 (19). Only a fraction of imported cases are generally detected at the destination (70; 27). According to the estimates proposed in Ref. (71), we stratify the detection capacity of countries into three groups: high, medium and low surveillance capacity according to the Global Health Security Index (72), and assume asymptomatic infections are never detected. The model calibration does not consider correlated importations (i.e., family travel) and assumes that travel probabilities are homogeneous across all individuals in each subpopulation. We perform for each state and country an additional ABC rejection analysis using as evidence the weekly reported deaths in the time window starting on March 22, 2020 through June 27, 2020. A full description of the model is provided in the SM file.

## Data Availability

Proprietary airline data are commercially available from Official Aviation Guide (OAG) and IATA databases. The GLEAM model is publicly available at http://www.gleamviz.org/.

## Acknowledgements

A.V., M.E.H., N.E.D., and I.M.L acknowledge support from NIH-R56AI148284 award. S.M. acknowledges support from the EU H2020 MOOD project. C.G. and L.R. acknowledge support from the EU H2020 Icarus project. M.A., M.C.and A.V. acknowledge support from COVID Supplement CDC-HHS-6U01IP001137-01. M.C. and A.V. acknowledge support from Google Cloud and Google Cloud Research Credits program to fund this project. A.V. acknowledges support from the McGovern and the Chleck Foundation. The findings and conclusions in this study are those of the authors and do not necessarily represent the official position of the funding agencies, the National Institutes of Health, or the U.S. Department of Health and Human Services.

## Author Contributions

J.T.D., M.C., N.P. and A.V. designed research; M.C., J.T.D., N.P., M.A., C.G., M.L., S.M., A.P.P, K.M., L.R., K.S., C.V, X.X., M.E.H., I.M.L., and A.V. performed research; M.C., J.T.D., N.P., A.P.P., K.M. and A.V. analyzed data; and M.C., J.T.D., N.P., M.A., C.G., M.L., S.M., A.P.P, K.M., N.E.D., L.R., K.S., C.V, X.X., M.E.H., I.M.L., and A.V. wrote and edited the paper.

