## Supplementary Information for "Cryptic transmission of SARS-CoV-2 and the first COVID-19 wave in Europe and the United States"

**4 SARS-CoV-2 Introduction Statistics** **13**

**5 SARS-CoV-2 Seeding Networks** **14**

**6 Correlation Analysis** **19**

**7 Data** **23**

*These authors contribute equally to this work,

### 1 Model Description

**1.1 Global Epidemic and Mobility Model.** We use the Global Epidemic and Mobility model (GLEAM), a stochastic, spatial, epidemic model based on an age-structured, metapopulation approach that has been used and published previously (1; 2). In the model, the world is divided into over 3,200 geographic subpopulations constructed using a Voronoi tessellation of the Earth's surface. Subpopulations, centered around major transportation hubs (e.g., airports), consist of cells with a resolution of 15 x 15 arc minutes (approximately 25 x 25 kilometers). High resolution data are used to define the population of each cell (3). Other attributes of individual subpopulations, such as age specific contact patterns, health infrastructure, etc., are added according to available data (4; 5).

GLEAM integrates a human mobility layer, represented as a network, using both short-range (i.e., commuting) and long-range (i.e., flights) mobility data from the Offices of Statistics for 30 countries on 5 continents as well as the Official Aviation Guide (OAG) and IATA databases (updated in 2019) (6; 7). The air travel network consists of the daily passenger flows between airport pairs (origin and destination) worldwide mapped to their corresponding subpopulations. We define a worldwide homogeneous standard for GLEAM to overcome differences in the spatial resolution of the commuting data across different countries. Where information is not available, the short-range mobility layer is generated synthetically by relying on the "gravity law" or the more recent "radiation law" both calibrated using real data available (8). These approaches assume more frequent travel to nearby or closer subpopulations and less frequent travel to distant locations. In Fig. 1 we show a representation of the geographical resolution of the model for a few selected regions, both the long range and short range mobility networks, and the population structure at the global level.

Initial conditions are set specifying the number and location of individuals capable of transmitting the infection. GLEAM is then able to track over time the proportion of the population in each disease compartment for all subpopulations. At the start of each simulated day, travelers move to their destinations via the flight network. The probability of air travel changes from day to day, varies by age group, and can consider the effects of location specific airline traffic reductions. Short-range mobility (i.e., commuting) varies by disease status. Each full day is simulated using 12 distinct time steps, and this process is repeated for every simulated day. Individuals and their traveling patterns are tracked as shown in the flow diagram for the GLEAM algorithm (Fig. 2).

The combined population structure and mobility network create a synthetic world that is used to simulate the unfolding dynamics of the epidemic. The infection dynamics occur within each subpopulation. We adopt a classic $SLIR$ model in which individuals are classified into four compartments: susceptible, latent, infectious, or removed. Susceptible individuals become latent through interactions with infectious individuals. Latent individuals progress to the infectious stage at a rate inversely proportional to the latent period, and infectious individuals progress to the removed stage at a rate inversely proportional to the infectious period. During both the latent and infectious stages we assume that individuals are able to travel. Following the infectious period, individuals then progress into the removed compartment where they are no longer able to infect others, meaning they have either recovered, been hospitalized, or isolated. Individuals transition between compartments using stochastic binomial chain processes assuming parameter values from available literature that define the natural history of disease. In Table. 1 we report the parameter estimates used in the model. We estimate the number of deaths using the number of individuals in the removed compartment and assume the infection fatality ratio has a uniform prior from $0.4\% - 2\%$ and is age-stratified proportional to the values reported in Verity et al. (9).

Once the mobility data layers and the disease dynamics are defined, the number of individuals in each compartment $m$, age bracket $i$, and subpopulation $j$ follows a discrete and stochastic dynamical equation that reads as

$$X_j^{[m,i]}(t + \Delta t) - X_j^{[m,i]}(t) = \Delta X_j^{[m,i]} + \Omega_j([m,i]) \quad (1)$$

where the term, $\Delta X_j^{[m,i]}$, represents the change due to the compartment transitions induced by the

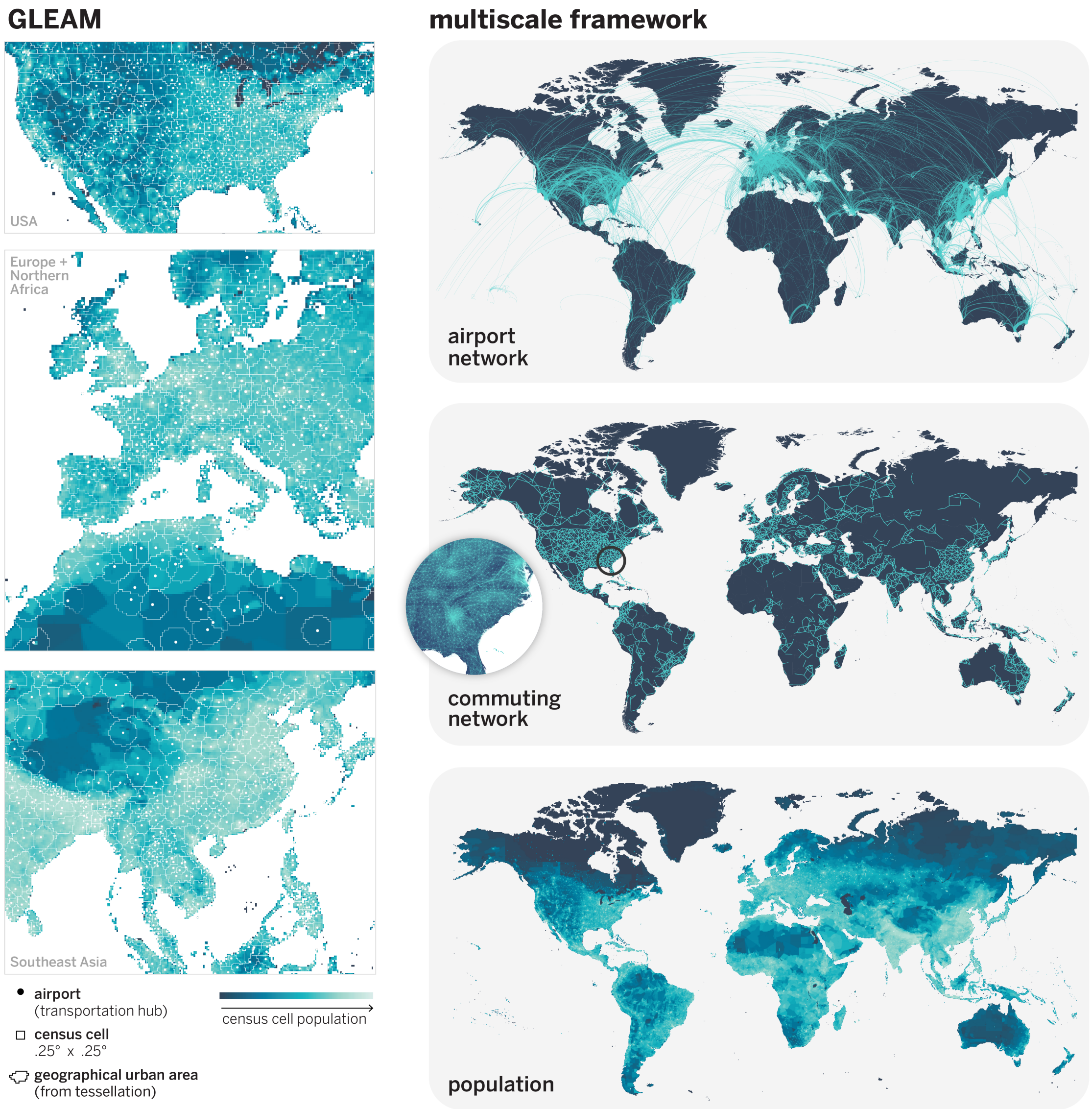

Figure 1: Schematic representation of GLEAM. (left) The subpopulation structure for selected regions. Subpopulations are geographic regions, formed from the Voronoi tessellation centered around airports. They are comprised of census cells that are approximately 25km x 25km. (right) Diagrams of the multiple mobility networks and population layer (from top to bottom): (1) the origin-destination airport network (long range mobility network), (2) the commuting network (short-range mobility network), (3) the population layer showing the population size of census cells.

disease dynamics and the transport operator, $\Omega_j([m,i])$, represents the variations due to the traveling and mobility of individuals. The latter operator takes into account the long-range airline mobility and defines the minimal time scale of integration as 1 day. The mobility due to the commuting flows is taken into account by defining effective force of infections by using a time scale separation approximation as detailed in Ref. (1). The $\Delta X_j^{[m,i]}$ is defined as the sum over all of the transitions into and out of disease compartment $m$ of individuals in age group $i$ ($[m,i]$). The operator $\mathcal{D}_j([m,i],[n,i])$ represents the number of transitions from $[m,i]$ to $[n,i]$ during the time interval $\Delta t$ and each element of this operator is a random variable extracted from a multinomial distribution. The change $\Delta X_j^{[m,i]}$ of a compartment $[m,i]$ in this

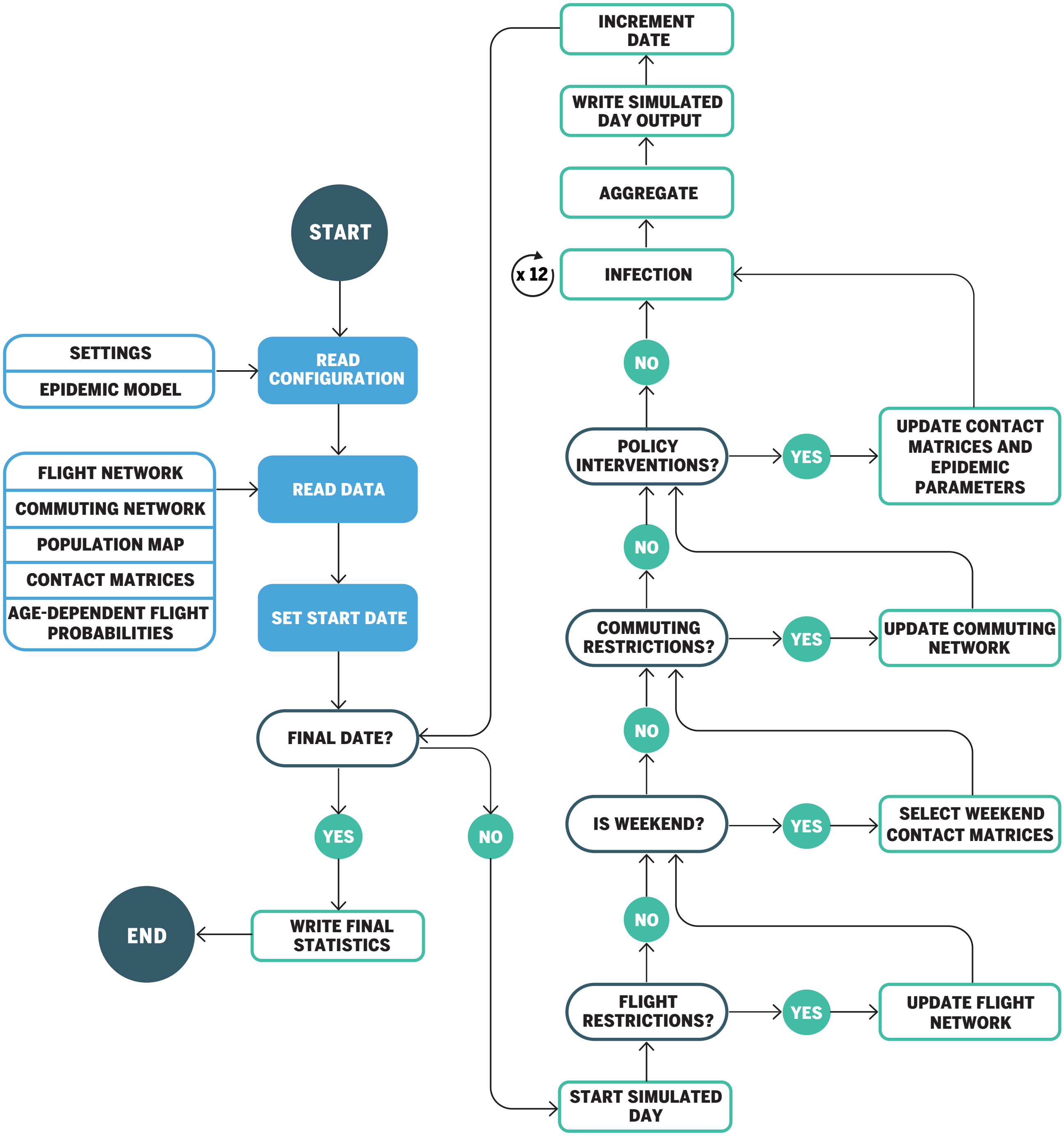

Figure 2: Flow diagram of GLEAM's algorithm

| Parameters | Range | Ref. |
|---|---|---|
| Latent period (mean) | [4, 7] days | (10) |
| Infectious period (mean) | [2, 4] days | (11) |
| Days until recovery | [10, 14] days | (11; 9) |
| Generation time | [6, 8] days | (12; 13) |

Table 1: Summary of parameter ranges explored in the sensitivity analysis. Reference parameters are reported in the main text

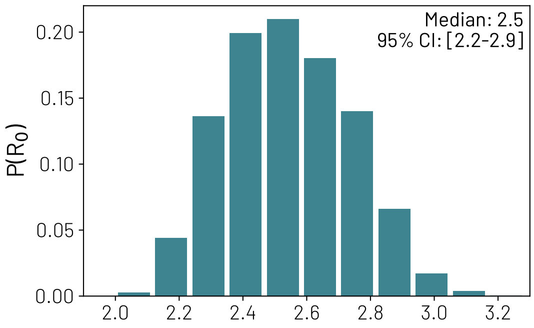

Figure 3: Posterior distribution of the reproductive number in China in the absence of mitigation policies.

time interval is given by a sum over all random variables $\{\mathcal{D}_j([m,i],[n,i])\}$ as follows

$$\Delta X_j^{[m,i]} = \sum_{[n,i]} \{-\mathcal{D}_j([m,i],[n,i]) + \mathcal{D}_j([n,i],[m,i])\}\,. \tag{2}$$

As a concrete example let us consider the evolution of the latent compartment. Individuals in age group $i$ of subpopulation $j$ can either transition into the Latent compartment ($L_j^i$) from the susceptible compartment ($S_j^i$) or transition out the Latent compartment into Infectious ($I_j^i$). The elements of the operator acting on $L_j^i$, are extracted from the binomial distributions: $Pr^{Bin}(L_j^i(t), p_{L_j^i \to I_j^i})$ and $Pr^{Bin}(S_j^i(t), p_{S_j^i \to L_j^i})$, where $p_{L_j^i \to I_j^i}$ and $p_{S_j^i \to L_j^i}$ are the transition probabilities from the latent state to the infectious state and from susceptible to the latent state, respectively. We assume a memoryless, discrete, stochastic transition processes. The probability $p_{S_j^i \to L_j^i}$ is the force of infection and it is determined by commuting flows, pattern of interactions as encoded in the age structured contact patterns, and the local non-pharmaceutical interventions. We consider individuals divided into 10 age groups: [0-9, 10-19, 20-24, 25-29, 30-39, 40-49, 50-59, 60-69, 70-79, 80+]. The contacts matrix $\mathbf{C}$ considers interactions in four specific social settings: contacts at school ($\mathbf{C}_{school}$), workplace ($\mathbf{C}_{work}$), home ($\mathbf{C}_{home}$), and in the general community ($\mathbf{C}_{community}$). Therefore, in general the contacts matrix is a linear combination of the four contributions according to the contact reductions in different locations $\mathbf{C} = \sum_s \omega_s \mathbf{C}_s$, where $\omega_s$ indicates the number of contacts per setting, and $s$ indicates the different settings mentioned before. The baseline $\omega_s$ and $\mathbf{C}_s$ values for each specific country are from Ref. (4). *For the sake of space we refer the reader to Ref. (1) where the analytical framework used in the model is reported in detail.*

**1.2 Interventions Timeline.** In order to realistically depict the evolution of the epidemic, a comprehensive set of policy interventions is applied to modify disease transmissibility and population mobility. On January 15, partial international travel reductions (from 10% to 40%) are applied for individuals traveling to/from China. Between January 23 and 28, flight and commuting reductions are applied to Wuhan and other subpopulations in the Hubei province to enforce government-mandated quarantines.

In addition, on January 25, commuting reductions are applied also to all other subpopulations in mainland China. To do so, we collected daily travel data starting January 1, 2020 until February 25, 2020 from the Baidu Qianxi platform (14), which provides three mobility indices (i.e., inflow index, outflow index, and intra-city index). The indices are proxies for the number of travelers moving in, out of, and inside a city, respectively. We extracted the mobility outflow index of 27 provinces and 4 municipalities for the year 2020 and the previous year (with the same lunar date), and then mapped all provinces and municipalities to the metapopulation structure of the model to estimate the travel flow changes during the epidemic where the travel reduction can be estimated as $1 - \frac{I_{cur}}{I_{pre}}$ , where $I_{cur}$ and $I_{pre}$ are the mobility outflow index of year 2020 and previous year on the same lunar date, respectively.

On February 1, due to the increasing amount of restrictions implemented by various countries and airlines (15; 16; 17; 18; 19; 20), stronger travel reductions are applied between mainland China and the rest of the world. We use actual worldwide (both international and domestic) origin-destination traffic data from the OAG database to quantify travel reductions. We also apply case detection based on travel history and additional travel bans across pairs of countries according to the Oxford COVID-19 Government Response Tracker (OxCGRT) (21). We account as well for the intra-country mobility and contacts reduction in workplaces and social settings (22) using the COVID-19 Community Mobility reports obtained from Google (23).

From mid-March 2020 all around the world, countries started to close schools as a means to slow the spread of COVID-19. We use the timeline of school closures provided by OxCGRT (21). As our model considers contact matrices for different settings, namely households, schools, workplaces and community contacts (4; 5), we quantify the decrease in contacts that individuals have in each of these environments. To implement school closures in the United States and the rest of the countries we follow (24) where authors study the effects of school closure in the context of seasonal influenza epidemics. According to the date when schools were closed in the different states/countries we consider a reduction of contacts in all individuals attending an educational institution (21). In the United States, Spain, and Italy, this intervention was applied at state/region level and for the rest of the European countries analyzed it was applied at country level.

Following school closures, most US states and European countries issued *stay-at-home* orders. In this case, we consider that only contacts in the household and essential workplaces were available. Using the COVID-19 Community Mobility reports (23) we compute the relative reduction on the number of contacts in workplaces and community interactions as well as the relative reduction in the intra-country mobility. We used data at the state or regional level for the United States, Italy, and Spain starting on February 15, 2020 and at the country level for all other countries available. For countries where we do not have mobility reports available we assume that on the date that schools closed there is a reduction in mobility of 50%, and an 100% reduction when there is a *stay-at-home* order. When the interventions are relaxed the mobility reduction is relaxed accordingly.

From the Google mobility reports we use the field `workplaces percent change from baseline` to infer contacts reductions in workplaces and the field `retail and recreation percent change from baseline` to infer contacts reductions in the general community setting. The Google mobility report provides the percentage change $r_l(t)$ on day $t$ of total visitors to specific locations $s$ with respect to a pre-pandemic baseline calculated as the median value, for the corresponding day of the week, during a 5 weeks period from January 3 until February 6, 2020. We turn this quantity into a rescaling factor for contacts such as $\omega_s(t) = \omega_s(1 + r_l(t)/100)^2$, by considering that the number of potential contacts per location scales as the square of the the number of visitors. We also use the ordinal index `C1 School closing` from the Oxford Coronavirus Government Response Tracker to modulate contacts in schools and universities. The index ranges from a minimum of 0 (no measures) to a maximum of 3 (require closing all levels). Furthermore, all $\omega$ factors are multiplied (or set equal to in case of contacts at home) by setting-specific weights from Mistry et al. (4). Finally we explore different level of overall transmissibility reduction (0-30%, step 10%) due to the awareness of population and behavioral changes starting at the date of the state of the emergency in the US and EU countries.

### 2 Model Calibration

The model described is stochastic and outputs an ensemble of possible epidemic outcomes for each set of initial conditions. We seed the epidemic in Wuhan, China assuming a starting date between November 15 and December 1, 2019, with 20 initial infections (25). Given the doubling time of the epidemic, this might corresponds to the virus emerging in mid October to late November, 2019 (26; 27; 28; 29; 25). We simulate epidemic scenarios sampling reproductive numbers ($R_0$) from a uniform prior in the range 1.6 to 3.3 (step

0.01). We use an Approximate Bayesian Computation (ABC) Rejection Algorithm to sample a set of parameter points $\theta$ (for instance $R_0$) according to a prior distribution and simulates through the model the dataset $E'$. A distance measure $s(E', E)$ determines the difference between $E'$ and the evidence $E$ based on a given metric. If the generated $E'$ is outside a tolerance from the evidence $E$ (i.e., $s(E', E) > \epsilon$) the sampled parameter value is discarded. The sampled parameters that are accepted provide an estimate of the likelihood with respect to the evidence $E$ and allows us to calculate the posterior distribution $P(\theta, E)$. As evidence, $E$, we considered the cumulative number of SARS-CoV-2 cases internationally imported from China during the time window of January 12 to January 21, 2020. The distance measure at each date is the difference between the SARS-CoV-2 cumulative imported cases generated by the model and the evidence with a tolerance provided by the under-detection interval estimated in Ref. (30). More specifically, only a fraction of imported cases are detected at the destination (31). According to the estimates proposed in Ref. (32), we stratify the detection capacity of countries relative to Singapore into three groups: high, medium and low surveillance capacity according to the Global Health Security Index (33), and assume an overall detection capacity for Singapore varying from 30% to 100% of imported cases. We also account for a non detectable 40% rate of asymptomatic individuals (sensitivity analysis ranging from 35% to 50%) (34; 35). The rejection algorithm accepts only configurations that satisfy the distance measure every day considered in the above time interval. This approach allows us to calibrate the model by incorporating both the growth rate of importations and their magnitude, scaled according to the under-detection estimates. The detailed list of importation events used is provided in Table S1 of the supplementary materials of Ref. (36). Using the ABC calibration and the age-stratified contact matrices, the obtained posterior distribution $P(R_0 = x|E)$ for the basic reproductive number $R_0$ in China has a median of 2.5 [95% CI 2.2-2.9] (Fig. 3), with a median doubling time of 3.8 [95% CI $3.1 - 4.6$] days in the absence of mitigation policies, for an overall detection capacity in Singapore of 60%. The posterior of $R_0$ in China has small variations, yielding a median of 2.4 [95% CI 2.1-2.8] and 2.7 [95% CI 2.3-3.1] for an overall detection capacity in Singapore of 100% and 30% respectively.

To estimate the posterior distribution of the infection fatality ratio (IFR) and infection attack rate in each US state and European country, we use an additional ABC rejection approach using the weekly model-projected and reported deaths. Specifically, we consider the subset of realizations that are (i) consistent with the international importations from China up to January 21, 2020 (i.e., selected from the global model calibration) and (ii) show Italy as the first country, in the group under examination, to experience sustained local transmission (more details in section 3A). Then we estimate, for each realization in each state and country considered, the projected deaths from the removed compartment by considering an uniformly distributed IFR prior ranging from 0.4% to 2% that is age stratified proportional to the values estimated by Ref. (9). We also consider that the projected deaths are subject to a reporting delay uniformly distributed between $2 - 22$ days for both the US and Europe. As a distance measure, $s(E', E)$, for the ABC rejection algorithm we use the summary statistics provided by the the weighted mean absolute percentage error ($wMAPE$):

$$wMAPE = \frac{\sum_t |D_{proj}(t) - D_{surv}(t)|}{\sum_t D_{surv}(t)} * 100$$

where $D_{proj}$ corresponds to the delayed/shifted model-projected deaths ($D_{proj}$) and $D_{surv}$ to the surveillance data. We only consider the deaths that were reported between March 22, 2020 and June 27, 2020, and set a tolerance of 25%, keeping only the realizations with a $s(E', E) = wMAPE < 25\%$. Using this approach we generate estimates and credible intervals for the infection attack rates and IFRs in 36 US states and 20 European countries. In the main text we show the results of the calibration on the weekly deaths for four US states and four European countries. In Fig. 4 and Fig. 5 we show the projected weekly deaths with the reported values for all calibrated European countries and US states. We also include Tables 2 and 3 which report the infection attack rate, infection fatality ratio (IFR), and reproductive number ($R_0$) of each for each US state and European country, respectively.

In Fig. 6 we show the correlation between the weekly projected deaths and the reported values from

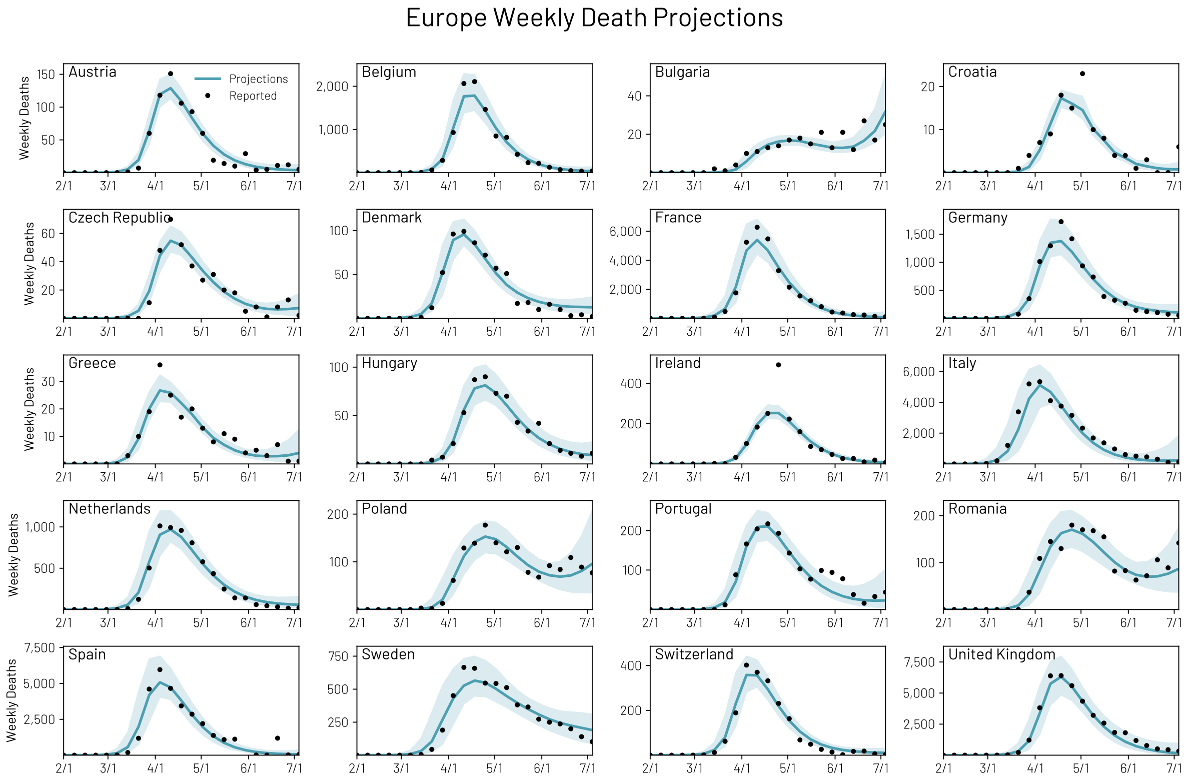

Figure 4: Projections of the weekly deaths for 20 European countries using the calibration reported in the main text. We report the median value and the 90% confidence interval.

| **Name** | Infection Attack Rate (%) | IFR (%) | $R_0$ |
|---|---|---|---|
| Austria | 1.16 [0.74, 2.85] | 0.81 [0.35, 1.23] | 2.61 [2.33, 2.83] |
| Belgium | 13.24 [8.50, 28.35] | 0.71 [0.33, 1.00] | 2.73 [2.34, 2.99] |
| Bulgaria | 0.98 [0.48, 2.35] | 1.06 [0.38, 1.63] | 2.66 [2.26, 2.80] |
| Croatia | 0.19 [0.12, 0.42] | 1.33 [0.57, 2.04] | 2.47 [2.24, 2.69] |
| Czech Republic | 0.46 [0.27, 1.22] | 0.86 [0.37, 1.31] | 2.59 [2.32, 2.83] |
| Denmark | 1.29 [0.82, 3.16] | 1.00 [0.41, 1.43] | 2.50 [2.24, 2.69] |
| France | 4.79 [3.38, 10.31] | 1.01 [0.47, 1.31] | 2.78 [2.47, 3.02] |
| Germany | 1.16 [0.73, 2.89] | 1.02 [0.42, 1.47] | 2.59 [2.34, 2.81] |
| Greece | 0.19 [0.10, 0.46] | 0.99 [0.40, 1.66] | 2.58 [2.34, 2.83] |
| Hungary | 0.80 [0.48, 2.02] | 0.87 [0.35, 1.27] | 2.62 [2.34, 2.85] |
| Ireland | 5.04 [3.20, 12.19] | 0.71 [0.30, 1.05] | 2.73 [2.41, 2.96] |
| Italy | 4.51 [3.13, 10.83] | 1.37 [0.63, 1.78] | 2.76 [2.38, 3.01] |
| Netherlands | 4.96 [3.13, 11.65] | 0.85 [0.37, 1.28] | 2.69 [2.37, 2.93] |
| Poland | 0.60 [0.30, 1.46] | 0.94 [0.39, 1.56] | 2.57 [2.32, 2.80] |
| Portugal | 1.56 [0.94, 3.67] | 1.07 [0.45, 1.53] | 2.69 [2.38, 2.92] |
| Romania | 1.07 [0.64, 2.53] | 0.94 [0.39, 1.33] | 2.69 [2.38, 2.95] |
| Spain | 7.30 [5.18, 14.79] | 1.09 [0.55, 1.38] | 2.76 [2.37, 3.02] |
| Sweden | 6.52 [3.98, 15.83] | 1.11 [0.42, 1.70] | 2.59 [2.31, 2.87] |
| Switzerland | 3.03 [2.00, 7.80] | 1.02 [0.42, 1.48] | 2.64 [2.34, 2.87] |
| United Kingdom | 6.68 [4.21, 15.05] | 0.97 [0.42, 1.39] | 2.72 [2.44, 2.93] |

Table 2: Model-estimated values for the infection attack rate by July 4, 2020, infection fatality ratio, and reproductive number ($R_0$) for the investigated European countries. We report the median values with the 90% CI

United States Weekly Death Projections

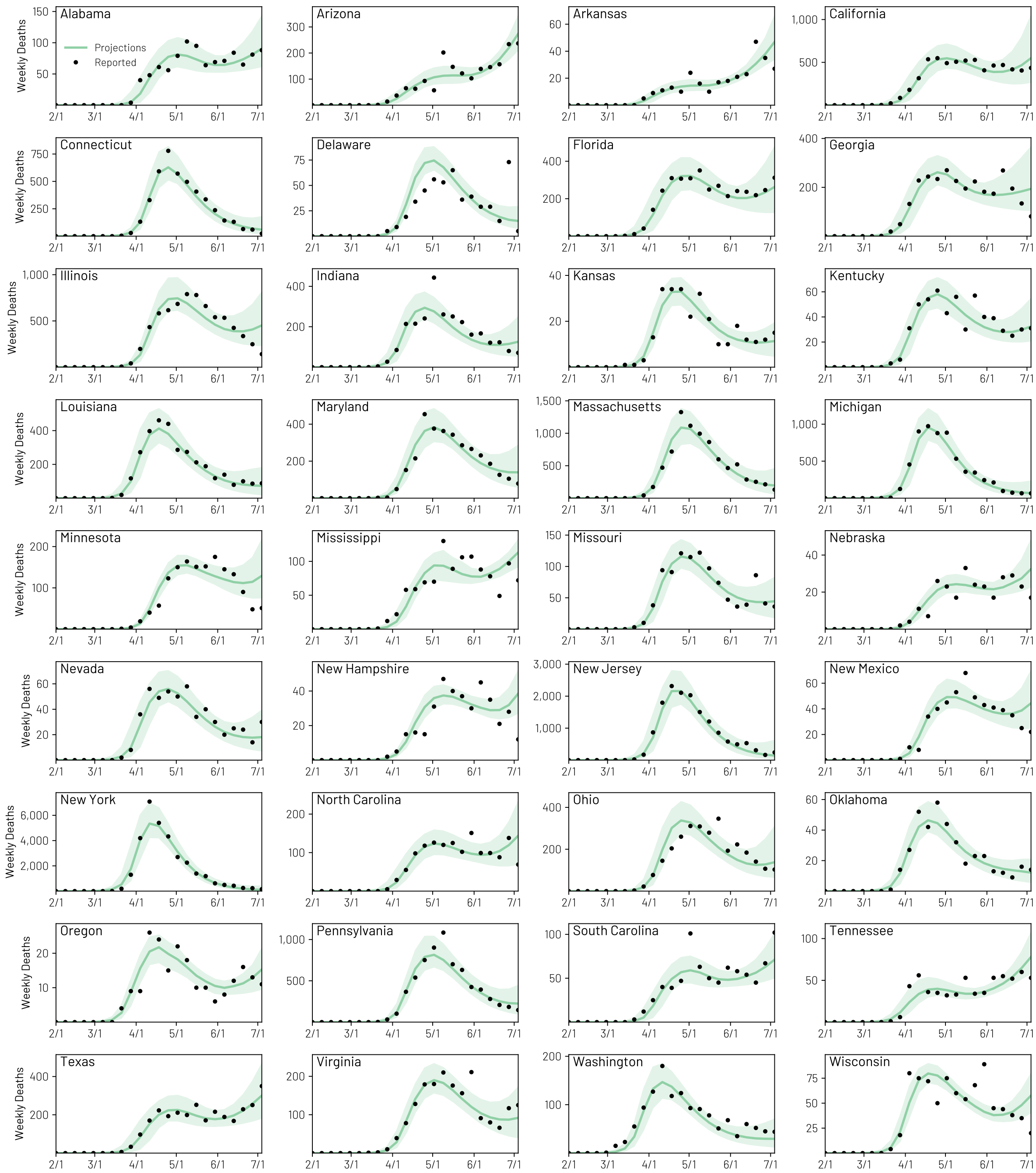

Figure 5: Projections of the weekly deaths for 36 US states using the calibration reported in the main text. We report the median value and the 90% CI.

| **Name** | Infection Attack Rate (%) | IFR (%) | $R_0$ |
|---|---|---|---|
| Alabama | 2.94 [1.68, 6.43] | 1.01 [0.44, 1.53] | 2.80 [2.51, 3.00] |
| Arizona | 5.95 [3.04, 12.96] | 0.95 [0.38, 1.59] | 2.60 [2.38, 2.76] |
| Arkansas | 1.52 [0.92, 4.07] | 1.12 [0.39, 1.61] | 2.64 [2.39, 2.93] |
| California | 3.08 [1.40, 6.97] | 0.83 [0.34, 1.40] | 2.48 [2.24, 2.69] |
| Connecticut | 9.44 [6.52, 19.60] | 1.30 [0.63, 1.60] | 2.80 [2.50, 3.04] |
| Delaware | 8.09 [5.37, 20.86] | 1.23 [0.48, 1.60] | 2.78 [2.43, 3.11] |
| Florida | 2.16 [1.29, 5.20] | 1.17 [0.51, 1.80] | 2.62 [2.36, 2.84] |
| Georgia | 4.46 [2.64, 9.99] | 0.89 [0.38, 1.29] | 2.71 [2.45, 2.95] |
| Illinois | 7.42 [4.41, 18.74] | 1.02 [0.43, 1.47] | 2.73 [2.41, 2.99] |
| Indiana | 4.83 [2.94, 11.55] | 1.07 [0.50, 1.49] | 2.80 [2.52, 3.02] |
| Kansas | 0.95 [0.57, 2.38] | 0.95 [0.41, 1.42] | 2.63 [2.37, 2.88] |
| Kentucky | 1.68 [1.08, 4.16] | 1.05 [0.43, 1.52] | 2.79 [2.50, 2.97] |
| Louisiana | 6.22 [4.26, 14.14] | 1.13 [0.50, 1.46] | 2.76 [2.40, 3.03] |
| Maryland | 6.53 [3.83, 14.77] | 1.03 [0.43, 1.48] | 2.74 [2.46, 2.97] |
| Massachusetts | 12.96 [7.80, 29.45] | 1.15 [0.49, 1.59] | 2.76 [2.52, 3.02] |
| Michigan | 6.39 [4.11, 13.79] | 1.03 [0.48, 1.46] | 2.72 [2.43, 3.02] |
| Minnesota | 5.76 [3.05, 13.29] | 0.94 [0.35, 1.43] | 2.76 [2.47, 2.94] |
| Mississippi | 5.19 [3.23, 10.67] | 1.16 [0.53, 1.54] | 2.76 [2.54, 2.98] |
| Missouri | 2.17 [1.38, 5.58] | 1.03 [0.42, 1.46] | 2.73 [2.45, 2.94] |
| Nebraska | 2.89 [1.75, 7.12] | 1.08 [0.44, 1.56] | 2.71 [2.53, 2.99] |
| Nevada | 2.74 [1.56, 6.68] | 0.91 [0.40, 1.41] | 2.62 [2.36, 2.86] |
| New Hampshire | 6.63 [3.53, 15.18] | 0.88 [0.39, 1.55] | 2.55 [2.37, 2.77] |
| New Jersey | 15.20 [10.22, 31.26] | 1.20 [0.60, 1.55] | 2.79 [2.49, 3.03] |
| New Mexico | 3.55 [2.25, 8.78] | 1.07 [0.45, 1.54] | 2.76 [2.42, 2.97] |
| New York | 13.37 [9.07, 26.72] | 1.14 [0.56, 1.50] | 2.78 [2.44, 3.02] |
| North Carolina | 2.81 [1.53, 6.45] | 0.97 [0.43, 1.50] | 2.70 [2.42, 2.92] |
| Ohio | 2.96 [1.67, 6.95] | 1.12 [0.46, 1.58] | 2.76 [2.52, 3.02] |
| Oklahoma | 1.04 [0.71, 2.74] | 1.04 [0.41, 1.44] | 2.61 [2.32, 2.86] |
| Oregon | 0.78 [0.48, 1.92] | 1.08 [0.44, 1.56] | 2.59 [2.37, 2.78] |
| Pennsylvania | 5.56 [3.46, 12.26] | 1.24 [0.57, 1.63] | 2.80 [2.43, 3.04] |
| South Carolina | 2.30 [1.36, 5.21] | 0.98 [0.43, 1.51] | 2.75 [2.49, 2.92] |
| Tennessee | 1.42 [0.90, 3.46] | 1.08 [0.42, 1.53] | 2.60 [2.45, 2.89] |
| Texas | 2.20 [1.20, 5.25] | 0.81 [0.36, 1.27] | 2.61 [2.33, 2.81] |
| Virginia | 2.81 [1.56, 6.09] | 0.96 [0.42, 1.46] | 2.71 [2.42, 2.92] |
| Washington | 1.84 [1.08, 4.28] | 0.92 [0.41, 1.44] | 2.64 [2.36, 2.88] |
| Wisconsin | 1.71 [1.21, 4.46] | 1.08 [0.41, 1.46] | 2.66 [2.51, 2.97] |

Table 3: Model-estimated values for the infection attack rate by July 4, 2020, infection fatality ratio, and reproductive number ($R_0$) for the investigated US states. We report the median values with the 95% CI

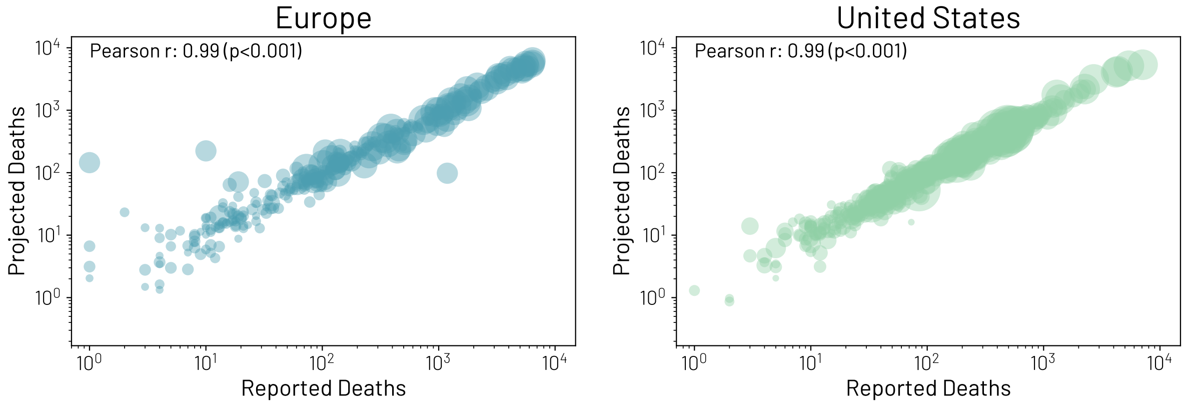

Figure 6: **Model calibration correlations.** The correlation between the median weekly projected deaths by the model reported in the main text and the weekly reported deaths in Europe (A) and the US (B). Each circle represents a weekly value for a single country/state and the size of the circle is proportional to the population size of the country/state.

surveillance data. We find a Pearson correlation coefficient of 0.99 ($p < 0.001$) from the results for both the US states and European countries. It is also important to stress that the calibration on reported weekly reported deaths is subject to the bias' of that data, for example: under-reporting, the use different definitions of COVID-19 deaths (e.g., some states/countries report both probable and confirmed deaths while others only report confirmed deaths), and outliers that are a result of states/countries reporting backlogged data on a single day.

Additionally, to analyze the stability of our selected list of states/countries from the calibration reported in the main text, we tested different tolerances of the $wMAPE$ scores. Increasing the tolerance to 30% adds 4 US states and 2 European countries (US: Utah, Maine, Colorado, Iowa; EU: Slovenia and Norway). Decreasing the tolerance to 20% removes 5 US states and 3 European country (US: Mississippi, Nebraska, New Hampshire, Oregon, Wisconsin; EU: Greece, Croatia, Bulgaria). However, the different tolerance values do not change the overall results using the calibration reported in the main text.

### 3 Sensitivity Analysis

**3.1 Unconstrained pandemic evolution realizations.** In Fig. 7 we show the rank distributions illustrating the probability, in our simulations, that each country started the local outbreak in a particular order $R$ (i.e., first, second, third etc.). While an initial start in the US and UK are the most likely scenarios in the ensemble (39% and 22% of simulations respectively), the empirical observations of case importation are also compatible with starts in other countries such as Germany, France, or Italy (13%, 10%, and 7% respectively). As a way to quantify and cluster the similarity of onset profiles, we compute and compare their cosine similarity. In particular, for each country, we create a vector where the $x_R$ component is the fraction of runs in which that country started the local outbreak in $R^{th}$ position. We then compute, for each pair, the cosine similarity building a similarity matrix. On the right side of Fig. 7 we show the correlation network with a threshold between pairs of 0.9. We use a community detection algorithm based on label propagation (37) that identifies three country clusters. These clusters represent country onset profiles that are considered to be similar to others in that group. The first group contains the US, UK, France, Germany, and Italy and these are all the countries among the first to have experienced the epidemic. The first confirmed cases in these countries were all reported within an eight day period. The second cluster instead is formed by countries such as Spain, Switzerland, Poland, and Portugal which are in the second group of countries to start observing local spreading of the virus. Spain acts as bridge with the first group. We find a third cluster, that includes all countries among the last to have experienced

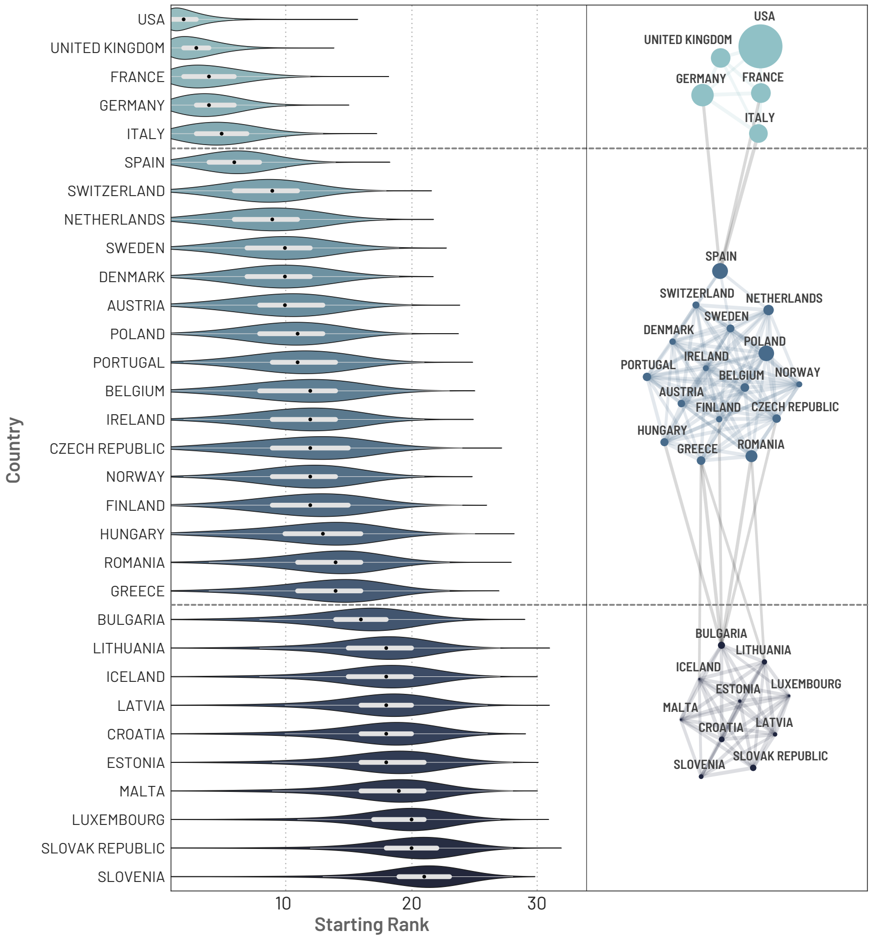

Figure 7: **Onset of local outbreaks in the selected ensemble.** On the left we show the distribution of starting ranks. The plot shows the probability that each country started the local outbreak in rank $R$ respect to the others. On the right we show the similarity network computed considering the cosine similarity of the starting rank distribution for each pair of countries. We threshold links showing just those equal or above to 0.9. The size of each node is proportional to the population of the respective country. The three clusters are identified via a community detection algorithm based on label propagation.

the epidemic such as Bulgaria, Iceland, and Lithuania. The first case detected within countries of this final cluster was on February 25, 2020 in Croatia (38), over a month after the first case was detected in Europe.

**3.2 Alternative distance measure for model calibration.** We further examine the robustness of the individual state and country results by performing an additional calibration that uses as distance measure in the ABC rejection algorithm the $s = ln(RSS)$, where the RSS is the residual sum of squares of the weekly deaths estimated by the model with respect to the weekly surveillance data. Similar to the calibration used in the main text, we analyze the time window from March 22, 2020, to June 27, 2020 and assume the same uniform prior distribution on the IFR range (0.4%-2%) and reporting delays $(2 - 22$ days). For each model realization that satisfies the global calibration, In we consider the empirical distribution of $P(s)$ and accept all the simulations for which $s$ is at a distance $\Delta < 0.66$, from the minimum value. This is equivalent to the typical information loss threshold between the model estimated deaths and the reported weekly incident deaths in information criteria. Using this selection, we

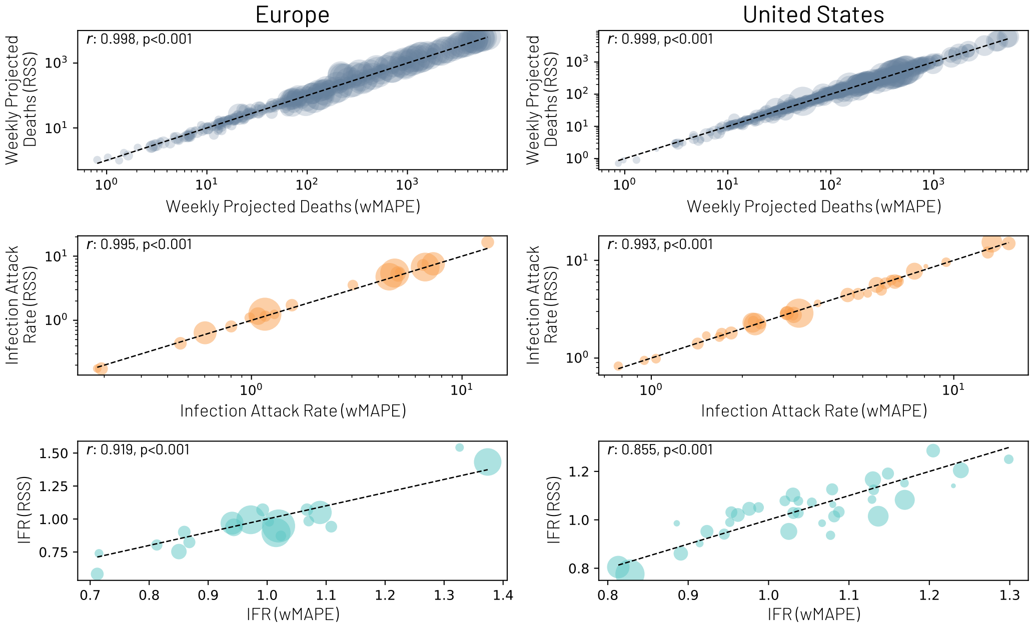

Figure 8: **Calibration comparison.** (A) The correlation between the weekly projected deaths using the ABC calibration in the main text (wMAPE) and the information theoretic calibration using the residual sum of squares distance metric (RSS). Each circle represents the weekly values for a single European country (left) or US state (right). (B) The correlation between the cumulative infection attack rates for each country/state as of July 4, 2020. (C) The correlation between the estimated IFRs for each calibration method. All circle sizes are proportional to the populations of each US state and European country.The dashed line in all figures represents the $y = x$ line.

confirm the results reported in the main text. In Fig. 8 we show the correlation of the weekly new deaths, infection attack rates as of July 4, 2020, and IFRs between the model calibrated using the $wMAPE$, ABC approach (from main text) and this information-theoretic approach.

### 4 SARS-CoV-2 Introduction Statistics

We record all introduction events through April 30, 2020 as described in the main text. We aggregate the observations from census areas to US states or European Countries (i.e., the targets). Then we construct a directed and weighted network in which importation sources link the target states/countries. The width of the link is the average share of importations from each source across all runs selected through April 30. Using these values, we build the chord diagrams shown in the manuscript. Since the weight of each link is the average across all runs of the normalized share of importation per run, the sum of incoming links for each target is one. To help the readability of the plots, we aggregated sources considering macro areas such as Europe and Asia. We keep the US (to isolate the national importations) and mainland China (as the epicenter of the pandemic) separate. All the other sources are grouped together and labeled "Others". More specifically, source countries of importations in are grouped as:

- **Asia**: Afghanistan, Armenia, Azerbaijan, Bahrain, Bangladesh, Brunei, Cambodia, Cyprus, India, Indonesia, Iran, Iraq, Israel, Japan, Jordan, Kazakhstan, Korea, Kuwait, Kyrgyzstan, Lao PDR, Lebanon, Malaysia, Maldives, Mongolia, Myanmar, Nepal, Oman, Pakistan, Philippines, Qatar, Saudi Arabia, Singapore, Sri Lanka, Taiwan, Tajikistan, Thailand, Turkey, United Arab Emirates, Uzbekistan, Vietnam, Yemen
- **China**: mainland China

- **Europe**: Albania, Austria, Belarus, Belgium, Bosnia and Herzegovina, Bulgaria, Croatia, Czech Republic, Denmark, Estonia, Finland, France, Germany, Gibraltar, Greece, Hungary, Iceland, Ireland, Isle of Man, Italy, Jersey, Kosovo, Latvia, Lithuania, Luxembourg, Macedonia, Malta, Moldova, Montenegro, Netherlands, Norway, Poland, Portugal, Romania, Russian Federation, Serbia, Slovak Republic, Slovenia, Spain, Sweden, Switzerland, Ukraine, United Kingdom
- **Others**: Algeria, Angola, Antigua and Barbuda, Argentina, Aruba, Australia, Bahamas, Barbados, Belize, Bermuda, Bolivia, Botswana, Brazil, British Virgin Islands, Burundi, Cameroon, Canada, Cape Verde, Caribbean Netherlands, Cayman Islands, Chile, Colombia, Congo, Cook Islands, Costa Rica, Cuba, Curaçao, Côte d'Ivoire, Djibouti, Dominica, Dominican Republic, Ecuador, Egypt, El Salvador, Equatorial Guinea, Ethiopia, Fiji, French Guiana, French Polynesia, Gambia, Ghana, Greenland, Grenada, Guadeloupe, Guatemala, Guinea, Guyana, Haiti, Honduras, Jamaica, Kenya, Liberia, Madagascar, Martinique, Mauritius, Mexico, Morocco, Mozambique, Namibia, New Zealand, Nicaragua, Nigeria, Palau, Panama, Paraguay, Peru, Rwanda, Samoa, Senegal, Seychelles, Sierra Leone, Somalia, South Africa, South Sudan, St-Barthélemy, St. Kitts and Nevis, St. Lucia, St. Maarten, St. Vincent and Grenadines, Sudan, Suriname, Tonga, Trinidad and Tobago, Tunisia, Turks and Caicos Islands, Uganda, Uruguay, Vanuatu, Venezuela, Zambia, Zanzibar, Zimbabwe
- **USA**: all the US states plus the US territories (American Samoa, Guam, Northern Mariana Islands, Puerto Rico, U.S. Virgin Islands)

In Table 4 we report the the share of introduction of SARS-Cov-2 infections for all European countries that experienced a local outbreak considering all the infections imported up to April 30, 2020. Compared with the seeding events networks (see below), the flows are radically different, especially for the first states and countries experiencing local transmission. The critical role of China before the travel restrictions of January 23, is replaced by a much larger fraction of introduction events of domestic or nearby countries origin.

In Table 5 we report the the share of introduction of SARS-Cov-2 infections for all US states considering all the infections imported up to April 30, 2020.

### 5 SARS-CoV-2 Seeding Networks

The importation networks are obtained as follows. As a first step, we track potential seeding events by air transportation (considering both individuals in the latent and infectious compartments) in any census areas of the US and Europe in all the runs selected. We then compute the day, in each run, in which the number of daily transitions from $S$ to $L$ is at least 10 in each state. In order words, we evaluate the date, in each run, when the state experienced the first local outbreak. We then track, in each run, the arrivals of latent and infectious individuals before or at the time of the local outbreak. From this standpoint, we build the seeding networks aggregating sources as described above.

In Table 6 we report the seeding share for all European countries considered (see also Fig. 9-B). Interestingly, China is the dominant seeding source for the first countries to have experienced the local outbreak such as Italy, UK, Germany, France and Spain. As we move down the list, towards countries that experienced a later start of the local outbreak, the share of seeding events from China rapidly decreases. Asia is a key source of infections for most of countries. For Denmark (57% [IQR 40% – 76%] ), Finland (56% [IQR 38% – 75%]), Sweden (56% [IQR 38% – 75%]), Austria (55% [IQR 36% – 75%]), Estonia (55% [IQR 36% – 75%]) and Switzerland (52% [IQR 33% – 73%]) the share of importations from Asian countries is above 50%. However, the role of European importation sources becomes more evident as we move clockwise in the plot thus looking at countries where the local outbreaks began in February. The range of importation shares goes from 10% [IQR 0% – 20%] in Italy to 87% [IQR 79% – 100%] in Slovak Republic. With the exceptions of the Netherlands (39% [IQR 20% – 57%]), Finland (37% [IQR

| State | Europe | China | Asia | USA | Others |
|---:|---:|---:|---:|---:|---:|
| Italy | 0.69 (0.6, 0.8) | <0.01 (<0.01, <0.01) | 0.21 (0.13, 0.28) | 0.06 (0.02, 0.06) | 0.04 (0.02, 0.05) |
| United Kingdom | 0.58 (0.48, 0.68) | <0.01 (<0.01, <0.01) | 0.27 (0.19, 0.35) | 0.08 (0.03, 0.09) | 0.07 (0.03, 0.08) |
| Germany | 0.53 (0.42, 0.64) | <0.01 (<0.01, <0.01) | 0.38 (0.27, 0.48) | 0.04 (0.02, 0.05) | 0.04 (0.02, 0.05) |
| France | 0.53 (0.42, 0.63) | <0.01 (<0.01, <0.01) | 0.27 (0.18, 0.35) | 0.06 (0.02, 0.07) | 0.15 (0.1, 0.18) |
| Spain | 0.84 (0.79, 0.91) | <0.01 (0.0, <0.01) | 0.10 (0.05, 0.12) | 0.04 (0.01, 0.04) | 0.02 (<0.01, 0.03) |
| Switzerland | 0.54 (0.42, 0.64) | <0.01 (0.0, <0.01) | 0.38 (0.27, 0.48) | 0.05 (0.02, 0.06) | 0.04 (0.01, 0.04) |
| Netherlands | 0.58 (0.47, 0.69) | <0.01 (0.0, <0.01) | 0.25 (0.16, 0.32) | 0.06 (0.02, 0.07) | 0.12 (0.06, 0.15) |
| Sweden | 0.67 (0.56, 0.77) | <0.01 (0.0, <0.01) | 0.28 (0.18, 0.37) | 0.04 (0.01, 0.04) | 0.02 (<0.01, 0.02) |
| Denmark | 0.54 (0.42, 0.65) | <0.01 (0.0, 0.0) | 0.39 (0.27, 0.5) | 0.05 (0.02, 0.05) | 0.03 (<0.01, 0.03) |
| Austria | 0.53 (0.42, 0.64) | <0.01 (0.0, <0.01) | 0.41 (0.3, 0.51) | 0.03 (0.01, 0.04) | 0.03 (0.01, 0.04) |
| Belgium | 0.57 (0.46, 0.69) | <0.01 (0.0, 0.0) | 0.33 (0.22, 0.43) | 0.05 (0.02, 0.06) | 0.04 (0.02, 0.05) |
| Poland | 0.73 (0.65, 0.83) | <0.01 (0.0, 0.0) | 0.21 (0.13, 0.28) | 0.03 (<0.01, 0.04) | 0.02 (<0.01, 0.03) |
| Portugal | 0.84 (0.79, 0.91) | <0.01 (0.0, 0.0) | 0.07 (0.04, 0.1) | 0.04 (0.01, 0.04) | 0.05 (0.01, 0.05) |
| Czech Republic | 0.62 (0.52, 0.72) | <0.01 (0.0, 0.0) | 0.31 (0.21, 0.4) | 0.04 (0.01, 0.04) | 0.04 (0.01, 0.04) |
| Ireland | 0.75 (0.67, 0.85) | <0.01 (0.0, 0.0) | 0.11 (0.06, 0.14) | 0.10 (0.04, 0.12) | 0.05 (0.01, 0.05) |
| Norway | 0.75 (0.67, 0.84) | <0.01 (0.0, 0.0) | 0.22 (0.13, 0.29) | 0.03 (<0.01, 0.03) | 0.01 (<0.01, 0.01) |
| Finland | 0.63 (0.53, 0.74) | <0.01 (0.0, 0.0) | 0.32 (0.22, 0.42) | 0.03 (<0.01, 0.03) | 0.02 (<0.01, 0.02) |
| Hungary | 0.70 (0.62, 0.81) | <0.01 (0.0, <0.01) | 0.23 (0.14, 0.29) | 0.04 (0.01, 0.05) | 0.03 (<0.01, 0.03) |
| Romania | 0.76 (0.69, 0.85) | <0.01 (0.0, 0.0) | 0.20 (0.11, 0.25) | 0.02 (<0.01, 0.03) | 0.02 (<0.01, 0.02) |
| Greece | 0.79 (0.73, 0.87) | <0.01 (0.0, 0.0) | 0.14 (0.08, 0.17) | 0.05 (0.01, 0.05) | 0.03 (<0.01, 0.03) |
| Bulgaria | 0.72 (0.65, 0.82) | <0.01 (0.0, 0.0) | 0.23 (0.14, 0.3) | 0.03 (<0.01, 0.03) | 0.02 (<0.01, 0.02) |
| Malta | 0.90 (0.87, 0.95) | <0.01 (0.0, 0.0) | 0.07 (0.03, 0.09) | 0.01 (<0.01, 0.01) | 0.02 (<0.01, 0.02) |
| Lithuania | 0.81 (0.75, 0.89) | <0.01 (0.0, 0.0) | 0.17 (0.1, 0.22) | 0.02 (0.0, 0.02) | <0.01 (0.0, <0.01) |
| Croatia | 0.73 (0.65, 0.82) | <0.01 (0.0, 0.0) | 0.18 (0.11, 0.23) | 0.04 (0.01, 0.05) | 0.05 (0.01, 0.06) |
| Latvia | 0.74 (0.67, 0.83) | <0.01 (0.0, 0.0) | 0.23 (0.14, 0.29) | 0.02 (0.0, 0.03) | 0.01 (0.0, 0.02) |
| Estonia | 0.65 (0.55, 0.76) | <0.01 (0.0, 0.0) | 0.31 (0.21, 0.41) | 0.03 (<0.01, 0.03) | <0.01 (0.0, 0.01) |
| Iceland | 0.69 (0.6, 0.82) | <0.01 (0.0, 0.0) | 0.07 (0.03, 0.09) | 0.16 (0.07, 0.2) | 0.08 (0.02, 0.1) |
| Luxembourg | 0.82 (0.76, 0.89) | <0.01 (0.0, 0.0) | 0.13 (0.07, 0.17) | 0.03 (<0.01, 0.04) | 0.02 (<0.01, 0.02) |
| Slovak Republic | 0.90 (0.86, 0.96) | <0.01 (0.0, 0.0) | 0.09 (0.03, 0.12) | <0.01 (0.0, 0.0) | <0.01 (0.0, <0.01) |
| Slovenia | 0.71 (0.62, 0.82) | <0.01 (0.0, 0.0) | 0.24 (0.14, 0.31) | 0.04 (0.0, 0.04) | 0.02 (0.0, 0.02) |

Table 4: Introduction of SARS-Cov-2 infections thorugh April 30. Sources are listed from the second column on. Targets are the European countries listed in the first column. Numbers are rounded to the second digit.

| State | USA | China | Asia | Europe | Others |
|---:|---:|---:|---:|---:|---:|
| California | 0.69 (0.6, 0.81) | <0.01 (<0.01, <0.01) | 0.11 (0.05, 0.15) | 0.05 (0.02, 0.07) | 0.14 (0.07, 0.19) |
| New York | 0.56 (0.43, 0.67) | <0.01 (<0.01, <0.01) | 0.11 (0.05, 0.15) | 0.14 (0.07, 0.19) | 0.19 (0.12, 0.23) |
| New Jersey | 0.56 (0.44, 0.68) | <0.01 (<0.01, <0.01) | 0.10 (0.05, 0.14) | 0.13 (0.06, 0.18) | 0.20 (0.12, 0.24) |
| Florida | 0.71 (0.62, 0.82) | <0.01 (0.0, <0.01) | 0.03 (0.01, 0.03) | 0.07 (0.03, 0.1) | 0.20 (0.11, 0.24) |
| Texas | 0.78 (0.71, 0.87) | <0.01 (0.0, <0.01) | 0.05 (0.02, 0.07) | 0.04 (0.02, 0.05) | 0.12 (0.06, 0.15) |
| Illinois | 0.71 (0.63, 0.82) | <0.01 (0.0, <0.01) | 0.07 (0.03, 0.1) | 0.06 (0.03, 0.08) | 0.15 (0.08, 0.18) |
| Washington | 0.81 (0.74, 0.89) | <0.01 (0.0, <0.01) | 0.06 (0.02, 0.08) | 0.03 (0.01, 0.04) | 0.10 (0.04, 0.12) |
| Massachusetts | 0.69 (0.6, 0.8) | <0.01 (0.0, <0.01) | 0.05 (0.02, 0.07) | 0.10 (0.04, 0.14) | 0.15 (0.09, 0.19) |
| Maryland | 0.71 (0.62, 0.82) | <0.01 (0.0, <0.01) | 0.08 (0.04, 0.11) | 0.08 (0.03, 0.1) | 0.12 (0.07, 0.15) |
| Nevada | 0.78 (0.71, 0.89) | <0.01 (0.0, 0.0) | 0.03 (<0.01, 0.03) | 0.04 (0.02, 0.05) | 0.15 (0.05, 0.2) |
| Virginia | 0.75 (0.67, 0.84) | <0.01 (0.0, <0.01) | 0.07 (0.03, 0.09) | 0.06 (0.03, 0.08) | 0.12 (0.07, 0.15) |
| Georgia | 0.78 (0.7, 0.86) | <0.01 (0.0, 0.0) | 0.07 (0.03, 0.09) | 0.05 (0.02, 0.06) | 0.10 (0.06, 0.13) |
| Arizona | 0.82 (0.77, 0.92) | <0.01 (0.0, <0.01) | 0.02 (<0.01, 0.03) | 0.02 (<0.01, 0.03) | 0.13 (0.05, 0.17) |
| Colorado | 0.83 (0.77, 0.9) | <0.01 (0.0, 0.0) | 0.02 (<0.01, 0.03) | 0.04 (0.01, 0.05) | 0.12 (0.06, 0.15) |
| Pennsylvania | 0.78 (0.72, 0.86) | <0.01 (0.0, 0.0) | 0.02 (<0.01, 0.03) | 0.05 (0.02, 0.07) | 0.14 (0.08, 0.18) |
| Ohio | 0.81 (0.76, 0.89) | <0.01 (0.0, <0.01) | 0.03 (<0.01, 0.03) | 0.04 (0.01, 0.05) | 0.12 (0.07, 0.15) |
| Connecticut | 0.64 (0.54, 0.76) | <0.01 (<0.01, <0.01) | 0.07 (0.03, 0.1) | 0.10 (0.04, 0.14) | 0.18 (0.11, 0.22) |
| Michigan | 0.79 (0.73, 0.87) | <0.01 (0.0, 0.0) | 0.03 (0.01, 0.04) | 0.05 (0.02, 0.06) | 0.13 (0.07, 0.16) |
| North Carolina | 0.81 (0.75, 0.89) | <0.01 (0.0, 0.0) | 0.03 (<0.01, 0.03) | 0.05 (0.02, 0.06) | 0.11 (0.06, 0.14) |
| Minnesota | 0.77 (0.69, 0.85) | <0.01 (0.0, 0.0) | 0.04 (0.02, 0.06) | 0.04 (0.01, 0.05) | 0.16 (0.09, 0.19) |
| Indiana | 0.78 (0.71, 0.87) | <0.01 (0.0, <0.01) | 0.04 (0.02, 0.05) | 0.05 (0.02, 0.06) | 0.13 (0.07, 0.16) |
| Oregon | 0.84 (0.79, 0.92) | <0.01 (0.0, 0.0) | 0.04 (0.01, 0.05) | 0.02 (<0.01, 0.03) | 0.10 (0.04, 0.12) |
| Utah | 0.86 (0.82, 0.93) | <0.01 (0.0, 0.0) | 0.03 (<0.01, 0.04) | 0.03 (<0.01, 0.04) | 0.08 (0.04, 0.11) |
| New Hampshire | 0.68 (0.58, 0.79) | <0.01 (0.0, <0.01) | 0.05 (0.02, 0.07) | 0.11 (0.05, 0.14) | 0.15 (0.09, 0.19) |
| Tennessee | 0.82 (0.76, 0.9) | <0.01 (0.0, 0.0) | 0.02 (<0.01, 0.03) | 0.04 (0.01, 0.05) | 0.12 (0.06, 0.15) |
| Missouri | 0.82 (0.76, 0.89) | <0.01 (0.0, 0.0) | 0.02 (<0.01, 0.03) | 0.03 (<0.01, 0.04) | 0.13 (0.07, 0.17) |
| Wisconsin | 0.85 (0.8, 0.92) | <0.01 (0.0, 0.0) | <0.01 (<0.01, 0.01) | 0.02 (<0.01, 0.03) | 0.12 (0.06, 0.16) |
| Louisiana | 0.84 (0.79, 0.91) | <0.01 (0.0, 0.0) | 0.02 (<0.01, 0.03) | 0.04 (0.01, 0.05) | 0.11 (0.05, 0.13) |
| South Carolina | 0.83 (0.78, 0.91) | <0.01 (0.0, 0.0) | 0.02 (<0.01, 0.03) | 0.04 (0.01, 0.06) | 0.10 (0.05, 0.13) |
| Kansas | 0.84 (0.79, 0.91) | <0.01 (0.0, 0.0) | 0.02 (<0.01, 0.02) | 0.03 (<0.01, 0.04) | 0.12 (0.06, 0.14) |
| Oklahoma | 0.83 (0.78, 0.91) | <0.01 (0.0, 0.0) | 0.03 (<0.01, 0.03) | 0.03 (<0.01, 0.04) | 0.11 (0.06, 0.14) |
| Kentucky | 0.82 (0.76, 0.9) | <0.01 (0.0, 0.0) | 0.03 (<0.01, 0.04) | 0.04 (0.01, 0.05) | 0.12 (0.06, 0.15) |
| Idaho | 0.88 (0.84, 0.94) | <0.01 (0.0, 0.0) | 0.02 (<0.01, 0.02) | 0.02 (<0.01, 0.02) | 0.08 (0.03, 0.11) |
| New Mexico | 0.89 (0.86, 0.95) | <0.01 (0.0, 0.0) | 0.01 (<0.01, 0.02) | 0.02 (<0.01, 0.03) | 0.07 (0.03, 0.09) |
| Iowa | 0.82 (0.76, 0.89) | <0.01 (0.0, 0.0) | 0.02 (<0.01, 0.03) | 0.03 (<0.01, 0.04) | 0.13 (0.07, 0.17) |
| Alabama | 0.70 (0.61, 0.83) | <0.01 (0.0, 0.0) | 0.18 (0.08, 0.24) | 0.03 (<0.01, 0.04) | 0.09 (0.04, 0.11) |
| Maine | 0.81 (0.75, 0.89) | <0.01 (0.0, <0.01) | 0.02 (<0.01, 0.03) | 0.04 (0.02, 0.06) | 0.12 (0.06, 0.16) |
| Alaska | 0.85 (0.8, 0.94) | <0.01 (0.0, 0.0) | 0.04 (<0.01, 0.05) | 0.02 (0.0, 0.02) | 0.09 (0.03, 0.12) |
| Nebraska | 0.82 (0.76, 0.9) | <0.01 (0.0, 0.0) | 0.02 (<0.01, 0.03) | 0.03 (<0.01, 0.04) | 0.13 (0.07, 0.17) |
| Rhode Island | 0.87 (0.83, 0.94) | <0.01 (0.0, 0.0) | <0.01 (0.0, 0.0) | 0.05 (<0.01, 0.06) | 0.08 (0.04, 0.11) |
| Montana | 0.89 (0.85, 0.95) | <0.01 (0.0, 0.0) | <0.01 (<0.01, 0.01) | 0.02 (<0.01, 0.02) | 0.09 (0.04, 0.11) |
| Arkansas | 0.84 (0.79, 0.91) | <0.01 (0.0, 0.0) | 0.03 (<0.01, 0.03) | 0.03 (<0.01, 0.04) | 0.10 (0.05, 0.13) |
| Delaware | 0.75 (0.67, 0.84) | <0.01 (0.0, 0.0) | 0.02 (<0.01, 0.03) | 0.06 (0.02, 0.08) | 0.17 (0.1, 0.21) |
| Mississippi | 0.85 (0.8, 0.92) | <0.01 (0.0, 0.0) | 0.01 (<0.01, 0.02) | 0.04 (<0.01, 0.05) | 0.10 (0.04, 0.13) |
| Vermont | 0.87 (0.83, 0.94) | <0.01 (0.0, 0.0) | 0.02 (0.0, 0.02) | 0.02 (0.0, 0.02) | 0.09 (0.04, 0.12) |
| West Virginia | 0.86 (0.81, 0.93) | <0.01 (0.0, 0.0) | 0.01 (<0.01, 0.01) | 0.03 (<0.01, 0.04) | 0.10 (0.04, 0.13) |
| Wyoming | 0.86 (0.81, 0.95) | <0.01 (0.0, 0.0) | 0.05 (<0.01, 0.06) | 0.02 (<0.01, 0.02) | 0.07 (0.02, 0.09) |
| North Dakota | 0.88 (0.85, 0.94) | <0.01 (0.0, 0.0) | <0.01 (0.0, 0.0) | <0.01 (0.0, <0.01) | 0.11 (0.05, 0.14) |
| South Dakota | 0.85 (0.8, 0.93) | <0.01 (0.0, 0.0) | <0.01 (0.0, 0.0) | 0.02 (0.0, 0.02) | 0.13 (0.06, 0.17) |

Table 5: Introduction of SARS-Cov-2 infections through April 30. Sources are listed from the second column on. Targets are the European countries listed in the first column. Numbers are rounded to the second digit.

| Country | Europe | China | Asia | USA | Others |
|---|---|---|---|---|---|
| Italy | 0.10 (0.0, 0.2) | 0.72 (0.5, 1.0) | 0.16 (0.0, 0.29) | <0.01 (0.0, 0.0) | <0.01 (0.0, 0.0) |
| United Kingdom | 0.15 (0.0, 0.23) | 0.52 (0.31, 0.71) | 0.28 (0.1, 0.43) | 0.02 (0.0, 0.0) | 0.04 (0.0, 0.0) |
| Germany | 0.22 (0.0, 0.33) | 0.31 (0.11, 0.5) | 0.43 (0.23, 0.62) | 0.02 (0.0, 0.0) | 0.02 (0.0, 0.0) |
| France | 0.22 (0.0, 0.33) | 0.41 (0.17, 0.62) | 0.33 (0.11, 0.5) | 0.02 (0.0, 0.0) | 0.02 (0.0, 0.0) |
| Spain | 0.55 (0.38, 0.75) | 0.15 (0.0, 0.2) | 0.26 (0.08, 0.4) | 0.02 (0.0, 0.0) | 0.02 (0.0, 0.0) |
| Switzerland | 0.32 (0.12, 0.5) | 0.09 (0.0, 0.11) | 0.52 (0.33, 0.73) | 0.03 (0.0, 0.0) | 0.03 (0.0, 0.0) |
| Netherlands | 0.39 (0.2, 0.57) | 0.12 (0.0, 0.14) | 0.42 (0.22, 0.6) | 0.03 (0.0, 0.0) | 0.04 (0.0, 0.0) |
| Sweden | 0.32 (0.15, 0.5) | 0.07 (0.0, 0.05) | 0.56 (0.38, 0.75) | 0.03 (0.0, 0.0) | 0.02 (0.0, 0.0) |
| Denmark | 0.33 (0.14, 0.5) | 0.06 (0.0, 0.0) | 0.57 (0.4, 0.76) | 0.03 (0.0, 0.0) | 0.02 (0.0, 0.0) |
| Austria | 0.34 (0.14, 0.5) | 0.07 (0.0, 0.04) | 0.55 (0.36, 0.75) | 0.02 (0.0, 0.0) | 0.02 (0.0, 0.0) |
| Belgium | 0.47 (0.25, 0.67) | 0.07 (0.0, 0.0) | 0.41 (0.2, 0.6) | 0.03 (0.0, 0.0) | 0.02 (0.0, 0.0) |
| Poland | 0.55 (0.35, 0.75) | 0.03 (0.0, 0.0) | 0.39 (0.18, 0.56) | 0.02 (0.0, 0.0) | 0.02 (0.0, 0.0) |
| Portugal | 0.70 (0.57, 0.89) | 0.05 (0.0, 0.0) | 0.19 (0.0, 0.3) | 0.02 (0.0, 0.0) | 0.04 (0.0, 0.0) |
| Czech Republic | 0.40 (0.18, 0.6) | 0.09 (0.0, 0.0) | 0.47 (0.25, 0.68) | 0.02 (0.0, 0.0) | 0.02 (0.0, 0.0) |
| Ireland | 0.56 (0.38, 0.76) | 0.05 (0.0, 0.0) | 0.25 (0.06, 0.38) | 0.07 (0.0, 0.1) | 0.07 (0.0, 0.09) |
| Norway | 0.47 (0.3, 0.64) | 0.01 (0.0, 0.0) | 0.48 (0.32, 0.67) | 0.03 (0.0, 0.0) | 0.02 (0.0, 0.0) |
| Finland | 0.37 (0.2, 0.53) | 0.03 (0.0, 0.0) | 0.56 (0.38, 0.75) | 0.02 (0.0, 0.0) | 0.02 (0.0, 0.0) |
| Hungary | 0.50 (0.29, 0.71) | 0.09 (0.0, 0.02) | 0.38 (0.17, 0.57) | 0.02 (0.0, 0.0) | 0.02 (0.0, 0.0) |
| Romania | 0.65 (0.5, 0.86) | 0.02 (0.0, 0.0) | 0.30 (0.11, 0.45) | 0.01 (0.0, 0.0) | 0.01 (0.0, 0.0) |
| Greece | 0.60 (0.43, 0.8) | 0.02 (0.0, 0.0) | 0.32 (0.12, 0.5) | 0.03 (0.0, 0.0) | 0.02 (0.0, 0.0) |
| Bulgaria | 0.60 (0.43, 0.8) | 0.01 (0.0, 0.0) | 0.36 (0.17, 0.5) | 0.02 (0.0, 0.0) | 0.01 (0.0, 0.0) |
| Malta | 0.79 (0.67, 1.0) | <0.01 (0.0, 0.0) | 0.18 (0.0, 0.25) | 0.01 (0.0, 0.0) | 0.02 (0.0, 0.0) |
| Lithuania | 0.69 (0.56, 0.87) | <0.01 (0.0, 0.0) | 0.29 (0.11, 0.42) | 0.01 (0.0, 0.0) | <0.01 (0.0, 0.0) |
| Croatia | 0.46 (0.25, 0.68) | <0.01 (0.0, 0.0) | 0.45 (0.21, 0.67) | 0.03 (0.0, 0.0) | 0.05 (0.0, 0.04) |
| Latvia | 0.57 (0.4, 0.76) | <0.01 (0.0, 0.0) | 0.39 (0.2, 0.56) | 0.02 (0.0, 0.0) | 0.01 (0.0, 0.0) |
| Estonia | 0.42 (0.21, 0.6) | <0.01 (0.0, 0.0) | 0.55 (0.36, 0.75) | 0.02 (0.0, 0.0) | <0.01 (0.0, 0.0) |
| Iceland | 0.61 (0.46, 0.8) | 0.01 (0.0, 0.0) | 0.16 (0.0, 0.23) | 0.14 (0.0, 0.2) | 0.08 (0.0, 0.1) |
| Luxembourg | 0.71 (0.58, 0.89) | <0.01 (0.0, 0.0) | 0.24 (0.08, 0.33) | 0.03 (0.0, 0.0) | 0.02 (0.0, 0.0) |
| Slovak Republic | 0.85 (0.79, 1.0) | <0.01 (0.0, 0.0) | 0.14 (0.0, 0.2) | <0.01 (0.0, 0.0) | <0.01 (0.0, 0.0) |
| Slovenia | 0.56 (0.4, 0.75) | <0.01 (0.0, 0.0) | 0.38 (0.18, 0.56) | 0.03 (0.0, 0.0) | 0.02 (0.0, 0.0) |

Table 6: Importation of seeding events. Sources are listed from the second column on. Targets are the countries listed in the first column. Numbers are rounded to the second digit.

| State | USA | China | Asia | Europe | Others |
|---|---|---|---|---|---|
| California | 0.09 (0.0, 0.14) | 0.74 (0.6, 1.0) | 0.13 (0.0, 0.2) | 0.01 (0.0, 0.0) | 0.03 (0.0, 0.0) |
| New York | 0.20 (0.0, 0.32) | 0.45 (0.15, 0.71) | 0.21 (0.0, 0.33) | 0.09 (0.0, 0.14) | 0.05 (0.0, 0.06) |
| New Jersey | 0.28 (0.11, 0.42) | 0.28 (0.07, 0.43) | 0.25 (0.1, 0.37) | 0.12 (0.0, 0.17) | 0.07 (0.0, 0.1) |
| Florida | 0.59 (0.42, 0.8) | 0.05 (0.0, 0.0) | 0.13 (0.0, 0.2) | 0.11 (0.0, 0.17) | 0.12 (0.0, 0.17) |
| Texas | 0.57 (0.39, 0.79) | 0.11 (0.0, 0.11) | 0.21 (0.0, 0.33) | 0.04 (0.0, 0.06) | 0.07 (0.0, 0.1) |
| Illinois | 0.46 (0.24, 0.68) | 0.19 (0.0, 0.23) | 0.22 (0.0, 0.34) | 0.06 (0.0, 0.08) | 0.07 (0.0, 0.1) |
| Washington | 0.56 (0.38, 0.78) | 0.11 (0.0, 0.11) | 0.23 (0.04, 0.36) | 0.03 (0.0, 0.03) | 0.07 (0.0, 0.08) |
| Massachusetts | 0.49 (0.29, 0.71) | 0.16 (0.0, 0.17) | 0.17 (0.0, 0.26) | 0.11 (0.0, 0.17) | 0.08 (0.0, 0.11) |
| Maryland | 0.48 (0.28, 0.7) | 0.11 (0.0, 0.09) | 0.25 (0.07, 0.37) | 0.09 (0.0, 0.14) | 0.07 (0.0, 0.09) |
| Nevada | 0.63 (0.47, 0.83) | 0.04 (0.0, 0.0) | 0.17 (0.0, 0.25) | 0.04 (0.0, 0.06) | 0.12 (0.0, 0.17) |
| Virginia | 0.55 (0.37, 0.75) | 0.08 (0.0, 0.06) | 0.23 (0.07, 0.33) | 0.08 (0.0, 0.12) | 0.07 (0.0, 0.08) |
| Georgia | 0.62 (0.46, 0.83) | 0.07 (0.0, 0.0) | 0.19 (<0.01, 0.29) | 0.06 (0.0, 0.08) | 0.06 (0.0, 0.08) |
| Arizona | 0.71 (0.57, 0.92) | 0.03 (0.0, <0.01) | 0.11 (<0.01, 0.15) | 0.03 (0.0, <0.01) | 0.12 (0.0, 0.17) |
| Colorado | 0.75 (0.64, 0.92) | 0.01 (0.0, 0.0) | 0.10 (0.0, 0.14) | 0.04 (0.0, 0.06) | 0.09 (0.0, 0.12) |
| Pennsylvania | 0.71 (0.56, 0.9) | 0.02 (0.0, 0.0) | 0.10 (0.0, 0.15) | 0.08 (0.0, 0.12) | 0.09 (0.0, 0.12) |
| Ohio | 0.71 (0.58, 0.9) | 0.03 (0.0, <0.01) | 0.13 (<0.01, 0.19) | 0.06 (0.0, 0.08) | 0.07 (0.0, 0.1) |
| Connecticut | 0.45 (0.24, 0.65) | 0.08 (0.01, 0.08) | 0.23 (0.09, 0.33) | 0.14 (0.04, 0.19) | 0.10 (0.01, 0.12) |
| Michigan | 0.63 (0.49, 0.85) | 0.08 (0.0, 0.0) | 0.16 (0.0, 0.23) | 0.07 (0.0, 0.1) | 0.07 (0.0, 0.1) |
| North Carolina | 0.68 (0.54, 0.88) | 0.06 (0.0, 0.0) | 0.13 (0.0, 0.18) | 0.07 (0.0, 0.09) | 0.07 (0.0, 0.09) |
| Minnesota | 0.68 (0.51, 0.87) | 0.01 (0.0, 0.0) | 0.16 (0.0, 0.25) | 0.05 (0.0, 0.07) | 0.10 (0.0, 0.13) |
| Indiana | 0.66 (0.5, 0.85) | 0.04 (0.0, 0.02) | 0.16 (0.04, 0.22) | 0.06 (0.0, 0.08) | 0.08 (0.0, 0.1) |
| Oregon | 0.73 (0.61, 0.9) | 0.03 (0.0, 0.0) | 0.15 (<0.01, 0.22) | 0.02 (0.0, 0.02) | 0.07 (0.0, 0.09) |
| Utah | 0.79 (0.67, 0.96) | <0.01 (0.0, 0.0) | 0.10 (0.0, 0.14) | 0.04 (0.0, 0.05) | 0.07 (0.0, 0.09) |
| New Hampshire | 0.53 (0.36, 0.72) | 0.09 (0.0, 0.06) | 0.16 (0.04, 0.23) | 0.13 (0.02, 0.19) | 0.09 (0.0, 0.12) |
| Tennessee | 0.74 (0.62, 0.92) | <0.01 (0.0, 0.0) | 0.11 (0.0, 0.15) | 0.05 (0.0, 0.07) | 0.09 (0.0, 0.12) |
| Missouri | 0.77 (0.67, 0.92) | <0.01 (0.0, 0.0) | 0.10 (0.0, 0.13) | 0.04 (0.0, 0.06) | 0.08 (0.0, 0.11) |
| Wisconsin | 0.83 (0.75, 0.98) | <0.01 (0.0, 0.0) | 0.04 (0.0, 0.03) | 0.03 (0.0, 0.01) | 0.09 (0.0, 0.13) |
| Louisiana | 0.78 (0.67, 0.94) | <0.01 (0.0, 0.0) | 0.08 (0.0, 0.11) | 0.05 (0.0, 0.08) | 0.08 (0.0, 0.11) |
| South Carolina | 0.77 (0.66, 0.93) | <0.01 (0.0, 0.0) | 0.10 (0.0, 0.13) | 0.06 (0.0, 0.08) | 0.06 (0.0, 0.08) |
| Kansas | 0.80 (0.7, 0.94) | <0.01 (0.0, 0.0) | 0.08 (0.0, 0.11) | 0.04 (0.0, 0.06) | 0.07 (0.0, 0.1) |
| Oklahoma | 0.79 (0.69, 0.96) | <0.01 (0.0, 0.0) | 0.08 (<0.01, 0.11) | 0.04 (0.0, 0.06) | 0.08 (<0.01, 0.11) |
| Kentucky | 0.77 (0.67, 0.92) | <0.01 (0.0, 0.0) | 0.10 (<0.01, 0.12) | 0.05 (0.0, 0.07) | 0.07 (0.0, 0.1) |
| Idaho | 0.84 (0.76, 0.98) | <0.01 (0.0, 0.0) | 0.07 (<0.01, 0.1) | 0.02 (0.0, <0.01) | 0.06 (0.0, 0.08) |
| New Mexico | 0.86 (0.79, 1.0) | <0.01 (0.0, 0.0) | 0.05 (0.0, 0.07) | 0.04 (0.0, 0.04) | 0.05 (0.0, 0.07) |
| Iowa | 0.78 (0.68, 0.94) | <0.01 (0.0, 0.0) | 0.08 (0.0, 0.1) | 0.04 (0.0, 0.05) | 0.09 (0.0, 0.13) |
| Alabama | 0.73 (0.6, 0.92) | <0.01 (0.0, 0.0) | 0.16 (<0.01, 0.22) | 0.05 (<0.01, 0.07) | 0.06 (<0.01, 0.08) |
| Maine | 0.77 (0.68, 0.91) | 0.02 (0.0, <0.01) | 0.07 (0.02, 0.08) | 0.07 (0.02, 0.09) | 0.08 (<0.01, 0.09) |
| Alaska | 0.78 (0.67, 0.94) | <0.01 (0.0, 0.0) | 0.12 (0.0, 0.17) | 0.02 (0.0, 0.0) | 0.08 (0.0, 0.12) |
| Nebraska | 0.81 (0.72, 0.94) | <0.01 (0.0, 0.0) | 0.06 (0.0, 0.07) | 0.04 (0.0, 0.05) | 0.09 (<0.01, 0.14) |
| Rhode Island | 0.87 (0.8, 1.0) | <0.01 (0.0, 0.0) | 0.02 (0.0, 0.0) | 0.06 (0.0, 0.08) | 0.05 (0.0, 0.07) |
| Montana | 0.90 (0.86, 1.0) | <0.01 (0.0, 0.0) | 0.01 (0.0, <0.01) | 0.02 (0.0, <0.01) | 0.07 (0.0, 0.1) |
| Arkansas | 0.83 (0.75, 0.97) | <0.01 (0.0, 0.0) | 0.06 (0.0, 0.08) | 0.05 (0.0, 0.07) | 0.06 (0.0, 0.09) |
| Delaware | 0.73 (0.63, 0.87) | <0.01 (0.0, 0.0) | 0.05 (0.01, 0.07) | 0.10 (0.03, 0.14) | 0.11 (0.03, 0.15) |
| Mississippi | 0.83 (0.75, 0.96) | <0.01 (0.0, 0.0) | 0.05 (0.0, 0.04) | 0.06 (0.0, 0.08) | 0.07 (0.0, 0.09) |
| Vermont | 0.87 (0.8, 1.0) | <0.01 (0.0, 0.0) | 0.05 (0.0, 0.06) | 0.03 (0.0, <0.01) | 0.05 (0.0, 0.07) |
| West Virginia | 0.85 (0.79, 0.96) | <0.01 (0.0, 0.0) | 0.03 (<0.01, 0.03) | 0.05 (<0.01, 0.05) | 0.07 (<0.01, 0.08) |
| Wyoming | 0.83 (0.75, 0.98) | <0.01 (0.0, 0.0) | 0.07 (<0.01, 0.09) | 0.02 (<0.01, <0.01) | 0.07 (<0.01, 0.09) |
| North Dakota | 0.91 (0.86, 1.0) | <0.01 (0.0, 0.0) | <0.01 (0.0, 0.0) | 0.01 (0.0, 0.0) | 0.08 (0.0, 0.11) |
| South Dakota | 0.88 (0.82, 1.0) | <0.01 (0.0, 0.0) | <0.01 (0.0, 0.0) | 0.02 (0.0, 0.0) | 0.09 (0.0, 0.13) |

Table 7: Importation of seeding events. Sources are listed from the second column on. Targets are the US states listed in the first column. Numbers are rounded to the second digit.

20% – 53%]), Austria (34% [IQR 14% – 50%]), Denmark (33% [IQR 14% – 50%]), Switzerland (32% [IQR 12% – 50%]), and Sweden (32% [IQR 15% – 50%]), all countries that experience a local onset of transmission after the first week of February are characterized by a share of European importations above or equal to 40%.

In Table 7 we report the seeding share for the US states. Within the US, while importations from mainland China contribute to early introductions of the virus, we find that other potential sources of importation play a key role in seeding the epidemic in different places. As shown in Fig. 9-A, the share of infection importations originating from Europe in California was nine times smaller than those in New York state (9% [IQR 0% – 14%]). Among the states for which the model estimates an early onset of local transmission before the third week of February (considering median values), European sources are statistically contributing 12% [IQR 0% – 17%] of SARS-CoV-2 importations for New Jersey, 11% [IQR 0% – 17%] for Florida, and only 4% [IQR 0% – 6%] for Texas. It is important to notice how, for countries in Europe, the US implemented additional travel advisories and restrictions a month later at the end February and early March. The share of importations from Asia is more significant for countries among the first to experience local outbreaks and becomes progressively smaller as we move clockwise in the plot. The range goes from 25% [IQR 10% – 37%] in New Jersey and 21% [IQR 0% – 33%] in New York to values smaller than 1% in North and South Dakota. As we mentioned above, the contribution from Asia is overall smaller than that of Europe. Interestingly, the domestic importations are, across the board, statistically relevant in seeding the epidemic in many states. Among the states for which we estimated a late onset of local transmission (second half of February), domestic sources account for 81% [IQR 72% – 94%] of the virus introductions in Nebraska, 86% [IQR 79% – 100%] in New Mexico, 83% [IQR 75% – 97%] in Arkansas, and 91% [IQR 86% – 100%] in North Dakota.

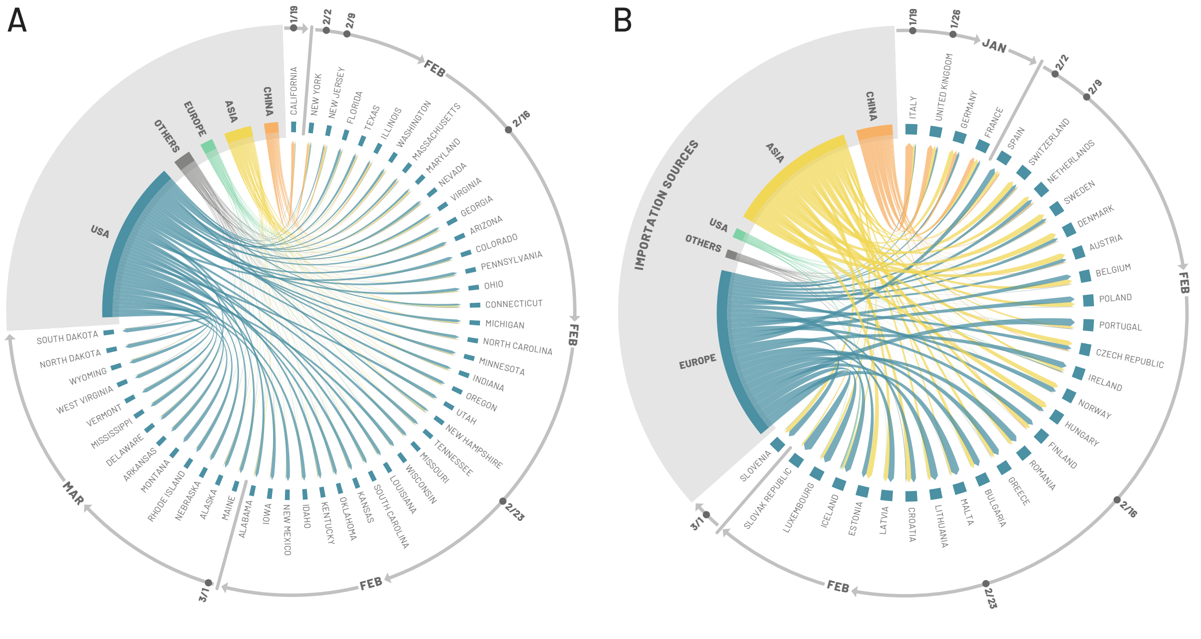

Figure 9: Share of importations of infections in all continental states (A) and in European countries (C) from US, China, Europe, Asia and all other countries before the start of the local outbreak. US states and European countries are ordered, clockwise, according to the start of the local outbreak.

### 6 Correlation Analysis

As mentioned and shown in the main text, during the early phases of the spreading, mobility plays a crucial role. In order to highlight this aspect, here we report the full correlation analysis between the

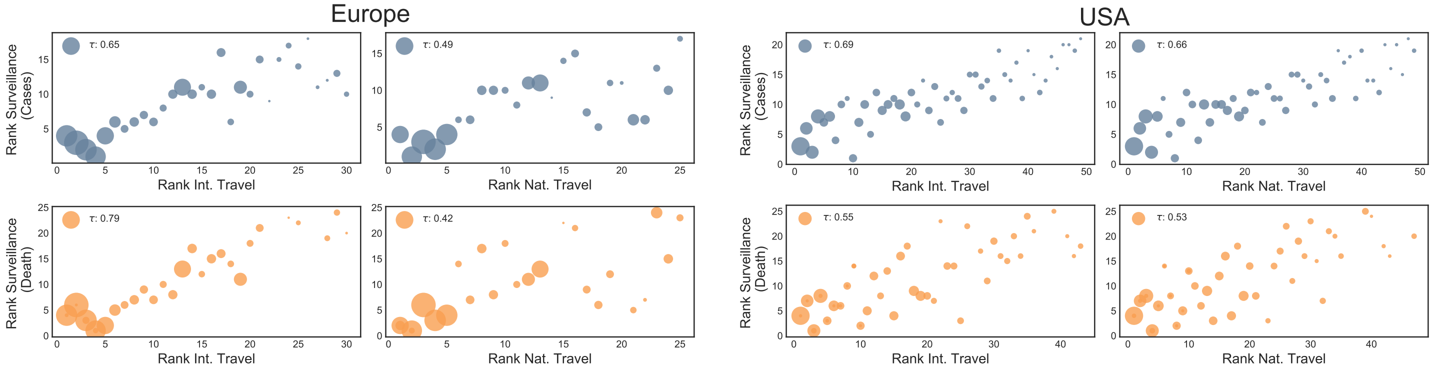

Figure 10: Correlation between the order in which states reached 100 confirmed cases (top row) or deaths (bottom row) and their International (left) or National (right) air traffic. The size of each state is assigned proportional to the population size. The first two columns refer to countries in Europe. The last two to the US states

real data and the mobility indicators. In particular, we compute the order in which states reached 100 cases/deaths in the real surveillance data and compare it with the order of European countries and US states according to their air traffic (considering both national and international travels). Note how the correlation plot in the main text considered as a mobility indicator the sum of the two types of traffic. In Fig. 10 we show the result reporting also the value of the Kendall's tau. In European countries both cases and deaths are highly correlated with international travels. The national flows are correlated by to a less extent. In US states, the rank of cases are more correlated to both international and national travels than deaths.

The countries and states that were the first to experience the outbreak, besides being hubs in the air transportation network, are also very populous. It is then natural to wonder how rankings based on population compare with respect to those based on air traffic. In Figure 11 we show the comparison. In particular, we order European countries and US states according to their population and density and to the epidemic indicators from surveillance (cases and deaths). We find high correlation levels with population ranks for both Europe and US states for both cases and deaths. It is interesting to note how those reported considering air travels are comparable or higher. The correlation for the number of deaths (bottom row) is lower with respect to the number of cases (top row) for US. Furthermore, it is interesting to notice how the correlations are even smaller when considering the population density, especially in the case of cases in Europe.

In Figure 12 we repeat the same analysis considering the model's projections. The correlations are comparable to the previous. Also in the model, population density is less correlated.

It is important to observe that air travel traffic, population and population densities are not independent indicators. Figure 13 highlights this observation. Particularly high is the correlation between air traffic and population. Also, population and population densities are well correlated (especially in USA) while air traffic and density are not. This is due, in part, to the many countries/states that, due to their location, see lots of traffic but are not very dense.

The correlations between rankings reported above have been computed by using the Kendall's tau (39) as implemented by the *scipy.stats* library (40). The metric is designed to compare the rankings obtained ordering items, states in our case, according to pairs of different quantities. The Kendall's tau is defined only in the case that the ranks have the same size. In case the two ranks have different size (i.e., some states did not yet go above a given threshold) the metric is applied to the common subset of the two.

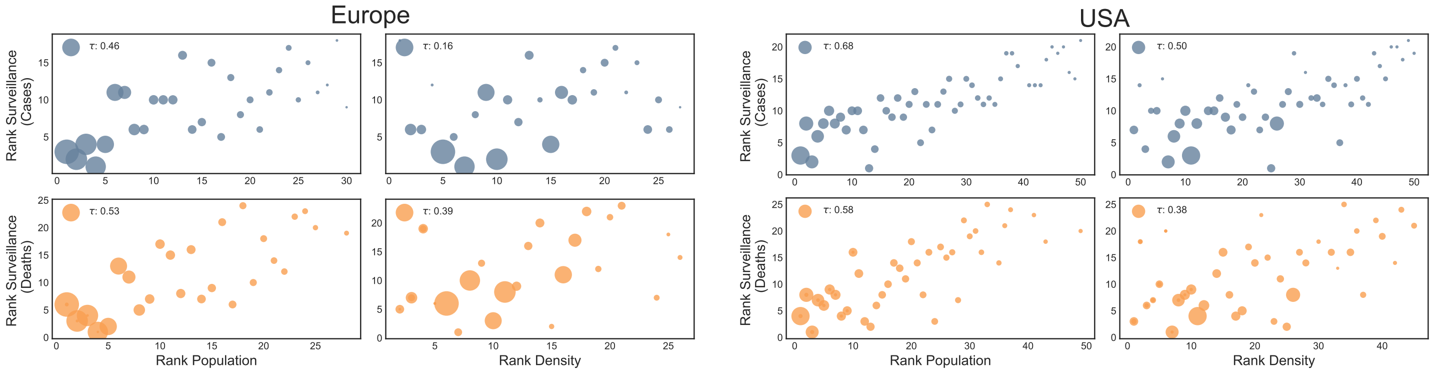

Figure 11: Correlation between the order in which states reached 100 confirmed cases (top row) or deaths (bottom row) and their population (left) or population density (right). The size of each state is assigned proportional to the population size. The first two columns refer to countries in Europe. The last two to the US states

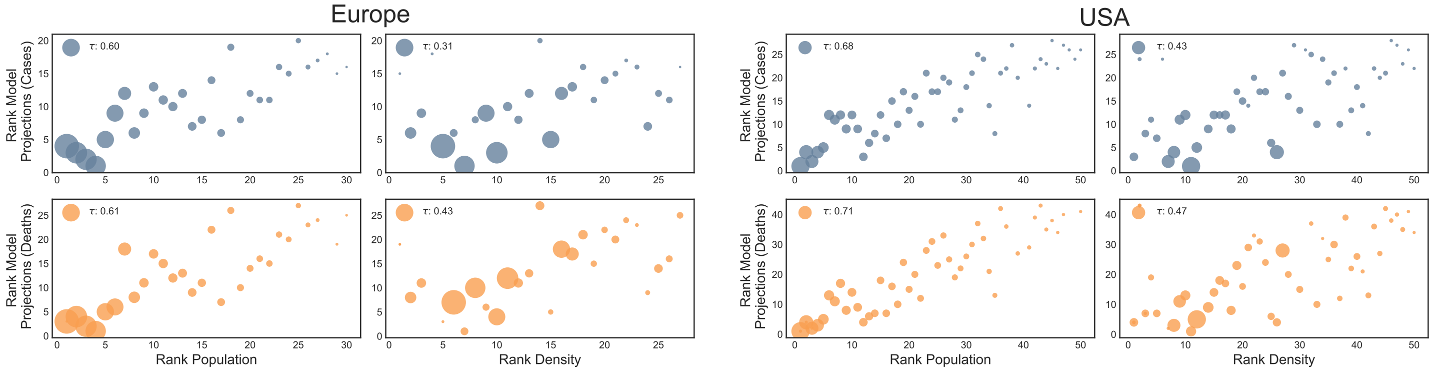

Figure 12: Correlation between the order in which states reached 100 confirmed cases (top row) or deaths (bottom row) according to the model and their population or population density. On the left we consider the case of European countries while on the right US states. The size of each country/state is assigned proportional to the population size

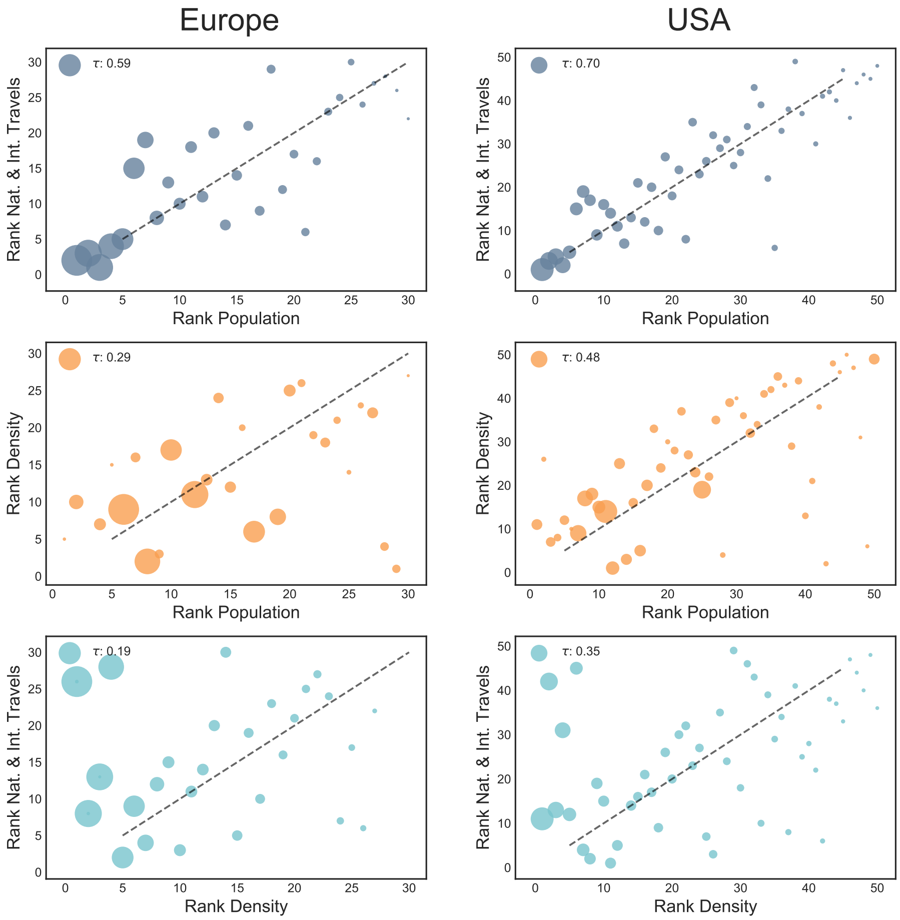

Figure 13: Correlation between travel, population and population density for the European countries (left) and continental US states (right)

| Country | Age | Study period | Prevalence (%) | Model AR (%) (median and 95%CI) | Reference |
|---|---|---|---|---|---|
| Denmark | 17-69 | 04/27/20-05/03/20 | 2.4 | 1.05 [0.67-3.03] | (42) |
| France | 0+ | 05/11/20-05/17/20 | 4.9 | 4.60 [3.13-11.05] | (43) |
| Czech Republic | 18-89 | 04/23/20-05/01/20 | 0.4 | 0.38 [0.22-1.02] | (44) |
| Portugal | 1+ | 05/21/20-07/08/20 | 2.7 | 1.60 [0.89-4.56] | (44) |
| Hungary | 14+ | 05/01/20-05/16/20 | 0.6 | 0.71 [0.42-1.99] | (45) |
| Spain | 0+ | 04/27/20-05/11/20 | 4.5 | 7.12 [4.95-16.05] | (46) |
| Italy | 0+ | 05/25/20-07/15/20 | 2.6 | 4.97 [3.06-17.43] | (47) |
| Sweden | 0-95 | 06/08/20-06/14/20 | 5.6 | 6.26 [3.52-16.86] | (48) |
| Netherlands | 18-72 | 05/11/20-05/18/20 | 5.6 | 4.67 [2.68-12.08] | (49) |
| Belgium | | 05/13/20 | 8.5 | 12.79 [7.90-31.25] | (50) |
| United Kingdom | | 05/24/20 | 6.78 | 6.44 [3.96-15.90] | (51) |
| **State/City** | **Age** | **Study period** | **Prevalence (%)** | **Model AR (%) (median and 95%CI)** | **Reference** |
| Los Angeles, CA | | 04/10/20-04/11/20 | 4.1 | 0.89 [0.27-2.91] | (52) |
| Connecticut | 0-65+ | 04/26/20-05/03/20 | 4.9 | 8.11 [5.77-19.26] | (53) |
| Louisiana | 0-65+ | 04/01/20-04/08/20 | 5.8 | 3.92 [2.59-11.57] | (53) |
| Minneapolis | 0-65+ | 04/30/20-05/12/20 | 2.4 | 3.87 [2.34-10.93] | (53) |
| Missouri | 0-65+ | 04/20/20-04/26/20 | 2.7 | 1.43 [0.91-4.14] | (53) |
| Philadelphia, PA | 0-65+ | 04/13/20-04/25/20 | 3.2 | 6.26 [1.35-21.11] | (53) |
| San Francisco | 0-65+ | 04/23/20-04/27/20 | 1 | 1.97 [0.32-7.79] | (53) |
| New York | 18+ | 04/19/20-04/28/20 | 14 | 12.98 [8.32-29.88] | (54) |
| New York City | 18+ | 04/19/20-04/28/20 | 22.7 | 19.78 [12.13-44.08] | (54) |

Table 8: Summary of the country and state level serological studies used for comparison against model estimates.

### 7 Data

**7.1 Epidemic surveillance data.** The surveillance data of the reported cases and deaths are taken from the John Hopkins University Coronavirus Resource Center (41).

**7.2 Model intervention data.** The model incorporates Google COVID-19 Community Mobility Reports data (23) to estimate, on the one hand, changes in mobility and, on the other hand, changes in contact patterns in workplaces and in the general community. Non-pharmaceutical interventions and other policy interventions are tracked using the Oxford Covid-19 Government Response Tracker (OxCGRT) (21). Lastly, reductions in air travel are computed by considering the percent change between the monthly origin-destination passenger flows between corresponding months in 2020 and 2019 (7). Implementation details are provided in Section 1.2.

**7.3 Serological data comparison.** We did an extensive literature search for serological studies performed from April-July 2020. In Fig. 5D, in the main text, we show the correlation between the estimated prevalence of SARS-CoV-2 antibodies and the model's estimated infection attack rate reported on the last date of that study. In Table 8 we report the prevalence values and study dates ranges for each serological survey considered along with our estimated infection attack rate.

[6] International Air Transportation Association; https://www.iata.org/.

[7] Official Aviation Guide; https://www.oag.com/.

[21] Hale, T., Webster, S., Petherick, A., Phillips, T. & Kira, B. Phylogenetic analysis of nCoV-2019 genomes. Oxford COVID-19 Government Response Tracker. Blavatnik School of Government (2020).

[22] Zhang, J. *et al.* Changes in contact patterns shape the dynamics of the COVID-19 outbreak in China. *Science* (2020). URL https://science.sciencemag.org/content/early/2020/04/28/science.abb8001. https://science.sciencemag.org/content/early/2020/04/28/science.abb8001.full.pdf.

[23] Google LLC. "Google COVID-19 Community Mobility Reports". https://www.google.com/covid19/mobility/.

[33] Global security index. https://www.ghsindex.org/.

[40] Scipy.org: Kendall Tau; https://docs.scipy.org/doc/scipy/reference/generated/scipy.stats.kendalltau.html.

[41] Johns Hopkins University Coronavirus Resource Center. https://coronavirus.jhu.edu/.
